# CXCR6+ T cells are required for CD3xCD20 Bispecific Antibody Efficacy in B cell Lymphomas

**DOI:** 10.1101/2024.07.02.24309792

**Authors:** Philipp Berning, Kazuya Fukasawa, Jahan Rahman, Lauren Melendez, Mythili Ketavarapu, Sneha Mitra, Chih-Yu Chen, Tetsuhiro Horie, Yasuhito Ishigaki, Monifa Douglas, Walter Ramos Amador, Daniela Hoehn, Ya-Hui Lin, Qi Gao, Mikhail Roshal, Mark D. Ewalt, Ahmet Dogan, Elisa DeStanchina, Benjamin Greenbaum, Christina S. Leslie, Gilles A. Salles, Lorenzo Falchi, Hans Guido Wendel, Santosha A. Vardhana

## Abstract

Activation of endogenous T cells using bispecific antibodies targeting CD3 and CD20 (BsAb) has emerged as a promising strategy for patients with B cell lymphomas. However, T cell features associated with treatment response or resistance are poorly understood. Here we identify an essential role for cytotoxic T cells in mediating response to BsAb in B cell lymphomas. In a disseminated B cell lymphoma model, BsAb showed substantial anti-tumor efficacy, which was dependent on activation and expansion of CD8+ T cells. Analysis of primary tumor samples from BsAb-treated patients confirmed that CD8+ T cell abundance was associated with durable remission and identified CXCR6+ cytotoxic T cells as associated with complete response. Cxcr6 knockout abrogated BsAb efficacy despite activation of CD4+ and CD8+ T cells. These results link BsAb efficacy to tumor resident cytotoxic T cells and suggest that enhancing chemokine-dependent T cell recruitment may augment therapeutic efficacy.

## INTRODUCTION

Follicular lymphomas (FL) and diffuse large B cell lymphomas (DLBCL) are the two most prevalent types of lymphoma. Response to standard immunochemotherapy approaches is variable and many patients do not achieve cure with standard approaches ^1, 2, 3^.

Therapeutic enhancement of anti-tumor T cell immunity has emerged as a clinically meaningful strategy in the treatment of both FL and DLBCL. While adoptive transfer of chimeric antigen receptor (CAR) T cells has shown substantial clinical activity, leading to approval of CAR T products for patients relapsed or refractory (R/R) FL or DLBCL ^4, 5, 6^, these treatments are only available at specialized treatment centers and are associated with a significant toxicity and morbidity profile. Bispecific antibodies (BsAb) are a novel class of pharmaceuticals that promote anti-tumor T cell immunity. These off-the-shelf drugs provide a mechanism for activating the endogenous T cell response and have shown significant activity in patients with various B cell lymphomas. At least four BsAb co-targeting CD3 and CD20 (CD3xC20), epcoritamab, glofitamab, mosunetuzumab, and odronextamab, have been developed with potent *in vitro* anti-lymphoma activity ^7, 8, 9, 10, 11^. In clinical trials, these agents demonstrated a significant rate of durable responses in heavily pre-treated patients with R/R FL or diffuse large B cell lymphoma (DLBCL), which led to regulatory approvals in both indications ^12, 13, 14, 15, 16^. This, in turn, promoted the rapid development of BsAb-based combinations with the objective of improving on single-agent results, and early data appear promising ^17, 18^.

Despite this progress, the mechanisms of BsAb response remain only partially understood. BsAb rely on recruitment of a patient’s endogenous T cells; as such, their efficacy may depend on both the composition and functional state of systemic and intratumoral T cell populations, as well as the capacity of BsAb to recruit and activate specific effector subsets. This is of particular importance in FL, where the microenvironment is known to be comprised of diverse immune populations, including several subsets of suppressive T and myeloid cells ^19, 20, 21^. Recent studies profiling patients on BsAb therapy have linked effective responses to T cell activation, proliferation, and cytotoxic gene expression ^8, 22, 23,24^. Despite these insights, current immune profiling approaches do not reliably distinguish patients with durable responses from those who experience progression ^25, 26, 27^. Direct functional dissection of CD4 versus CD8 roles in BsAb-mediated activity may be a crucial step in characterizing effective, long-lasting BsAb responses. To that end, we analyzed intratumoral T cell subpopulations in both a newly established Bcl2/Ezh2^Y646F^ dependent syngeneic mouse lymphoma model ^28^ treated with BsAb and in patients with B cell lymphomas receiving BsAb-based combinations and identified both CD8+ and CXCR6+ T cells as key drivers of an effective BsAb response *in vivo*.

## RESULTS

### CD3xCD20 BsAb promote tumor clearance in a disseminated Bcl2/Ezh2-driven B cell lymphoma model

To define the T cell programs that accompany BsAb-mediated tumor control *in vivo*, we established an immunocompetent, disseminated model of germinal center B-NHL via adoptive transfer of a previously described B cell line (termed tFL-P6) derived from an *Ezh2^Y641F^; Rosa26^LSL-BCL^*^2^*^-IRES-GFP^* mouse model ^28, 29^ (**Figure 1A)**. Following adoptive transfer, mice developed splenomegaly within 7 days and died within 20 days with histopathology demonstrating lymphomatous involvement. Treatment of mice 7 days after tumor implantation with a single dose of a murine CD3xCD20 BsAb (provided by Genentech) ^30^ significantly prolonged survival compared with mice treated with an isotype IgG control (median not reached vs 11 days; p<0.0001) (**Figure 1B)**.

**Figure 1.**
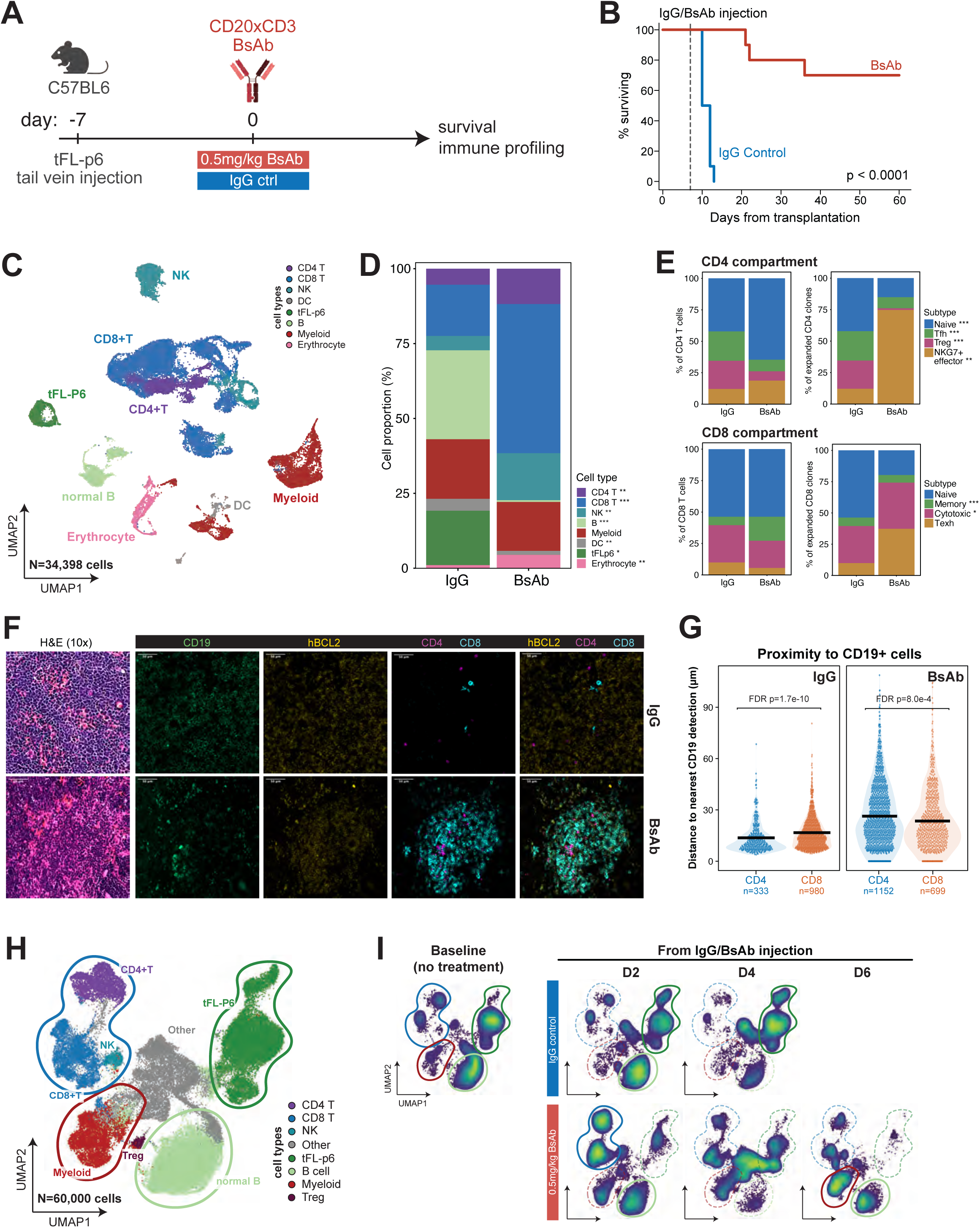
A CD3xCD20 bispecific antibody eradicates BCL2/Ezh2 lymphomas *in vivo*. (A–B) Experimental schema (A) and survival (B) of BCL2/Ezh2 tumor-bearing mice following BsAb treatment (n=10/group). P value determined by log-rank test. (C) UMAP embedding of the full single-cell dataset (N=34,398 cells; n=4 mice/group), colored by cell type. (D) Broad cell-type composition by treatment condition (IgG vs. BsAb) in tumor-bearing mice after 4 days of treatment (n=4 mice/group). Asterisks indicate differential abundance (BsAb vs. IgG) by propeller (moderated t-test on arcsine square-root–transformed proportions). *p < 0.05, **p < 0.01, ***p < 0.001. (E) CD4+ (top) and CD8+ (bottom) T-cell compartment composition among total cells (left) and clonally expanded cells (right) by treatment condition. Asterisks (total cells only) indicate differential abundance by propeller (moderated t-test on arcsine square-root–transformed proportions). *p < 0.05, **p < 0.01, ***p < 0.001 Expanded-clone composition is shown only descriptively. (F) Representative multiplexed immunofluorescence of spleens harvested 4 days after treatment with IgG control or BsAb, showing H&E (10x) and CD19, hBCL2, CD4, and CD8 staining, with composite overlays highlighting spatial proximity of T cells to tumor regions. (G) Multiplex immunofluorescence-based quantification of the distance from CD4+ and CD8+ T cells to the nearest CD19+ cell following IgG or BsAb treatment. Each point represents an individual cell detection; black bars indicate means. FDR-adjusted p values were determined using Wilcoxon rank-sum tests. (H) UMAP embedding of 60,000 downsampled events from spectral flow cytometry, equally pooled across all treatment conditions, showing major immune and tumor-cell populations. (I) Condition- and time-resolved UMAP density plots for IgG control (top) and BsAb (bottom) at baseline (day 0) and days 2, 4, and 6 after treatment.

To determine how BsAb remodel the tumor microenvironment to drive tumor regression, we performed paired CITE-seq and scTCR-seq on spleens from tumor-bearing mice treated with IgG isotype control or BsAb for 4 days (n = 4/arm). Louvain-based clustering resolved 8 major cell populations, including CD4+ T, CD8+ T, NK, B, myeloid, DC, tFLp6 cells, and erythrocytes **(Figure 1C** and **Supplementary Figure 1A**). BsAb treatment nearly eliminated malignant cells (19.6% to 0.06%; propeller, p = 0.013) and expanded CD4+ T cell, CD8 T cell, and NK cell populations (all p < 0.01; n=4 per arm) (**Figure 1D**). Sub-clustering of CD4+ and CD8+ T cells revealed 4 major subsets within both CD4+ (naïve, Tfh, Treg, and NKG7+ effector) and CD8+ (naïve, memory, exhausted, and cytotoxic) T cell states (**Supplementary Figure 1B,C).** The abundance of T cell subtypes shifted with treatment: within the CD4 compartment, BsAb treatment depleted Tfh and Treg cells and expanded naïve and NKG7⁺ effector cells, while among CD8+ T cells, memory cells increased from 7% to 20% (propeller, p < 0.005 for each; **Figure 1E**). Integrated scTCR analysis revealed that the majority of clonally expanded CD4+ T cells following BsAb therapy were NKG7+ effectors (75% vs. 11% with IgG), whereas expanded CD8+ T-cell clones comprised a mixture of cytotoxic and exhausted phenotypes, with exhausted (Texh) clones increasing from 9% to 36% (decriptive comparison due to low T cell recovery in IgG-treated mice; **Figure 1E**). Multiplexed immunofluorescence of spleens performed 4 days after BsAb administration confirmed remodeling of the tumor microenvironment with reduced tumor burden as indicated by a reduction in CD19/hBCL2 co-expressing cells, and substantial accumulation of T cells, the majority of which were CD8+ (**Figure 1F**). BsAb treated tumors also showed a redistribution of intratumoral T cells, with CD8+ T cells more proximal to tumors than CD4+ T cells, in contrast to control-treated tumors, in which CD4+ T cells were more tumor-proximal (**Figure 1G**).

To longitudinally evaluate the cellular changes associated with BsAb-mediated tumor control in this model, we treated mice bearing Bcl2/Ezh2 tumors with BsAb or isotype control and profiled single-cell suspensions of splenocytes at days 2, 4, and 6 after treatment using a 22-color multiparametric spectral cytometry panel (**Supplementary Table 1**). Unsupervised clustering of viable (Live/Dead-negative) single cells based on expression of 21 cell markers resolved eight major cell populations: malignant B cells (B220, CXCR5, GFP), normal B cells (B220, CXCR5), myeloid cells (CD11b), NK cells (NK1.1), CD8⁺ T cells (TCRb, CD8), CD4⁺ T cells (TCRb, CD4), regulatory T cells (TCRb, CD4, FoxP3), and an eighth population that could not be clearly distinguished based on the chosen panel (**Figure 1H, Supplementary Figure 1D**). BsAb-treated spleens over time showed a clear reduction in splenic tumor burden, with tFLp6 becoming almost undetectable within 2 days of BsAb treatment initiation while in control Ab-treated mice, tumor burden steadily progressed, requiring euthanasia before 6 days had elapsed (**Figure 1H and Supplementary Figure 1E**). In parallel, spleens from BsAb-treated mice exhibited substantial remodeling of non-malignant immune cell compartments, with a significant increase in both CD4+ and CD8+ T cells within 2 days of BsAb initiation and a subsequent increase in myeloid and NK cells (**Supplementary Figure 1F**).

Follicular lymphoma frequently involves not only secondary lymphoid organs including the spleen and LN but also the bone marrow, the latter of which is frequently the site of residual disease following B cell depleting therapies ^31^. To determine whether BsAb efficiently eradicated tFLp6 cells across tissues, we performed spectral flow cytometry on single-cell suspensions from the spleen, bone marrow, and lymph nodes of tumor-bearing mice at day 4 of IgG or BsAb treatment. Broad immune composition analysis showed that tFLp6 cells were primarily detectable in the spleen and bone marrow, with only minimal involvement of the lymph nodes (**Supplementary Figure 1G**). Following BsAb treatment and after normalization of the resolved populations, tFLp6 cells decreased from 20.2% to 0.045% in the spleen and from 29.8% to 0.129% in the bone marrow, accompanied by relative expansion of myeloid, CD8+ T cell, and NK cell populations (**Supplementary Figure 1G**). Analysis as a proportion of live cells similarly showed a strong trend toward reduced median tFLp6 abundance in the spleen (1.56% to 0.011%) and bone marrow (16.2% to 0.055%; p=0.057 for both), together with significant increases in NK cells in both tissues (spleen, 1.80% to 3.51%; bone marrow, 1.27% to 1.68%; both p < 0.05). Although tFLp6 cells were rare in the lymph nodes, BsAb treatment significantly reduced normal B cell abundance (44.10% to 20.15%) while increasing CD8+ T cells (15.85% to 33.95%) and NK cells (1.17% to 2.65%; all p < 0.05), indicating systemic immune remodeling despite the low local tumor burden (**Supplementary Figure1H**).

### CD3xCD20 BsAb remodel the tumor microenvironment to support cytotoxic T cell function

We next asked whether CD3xCD20 BsAb therapy preferentially engaged specific T cell subsets and what the impact of BsAb engagement was on T cell function. CD8+ and CD4+ T cells from spleens of tumor-bearing mice at 2, 4, and 6 days after treatment with IgG or BsAb were separated into subclusters based on expression of canonical markers (**Supplementary Figure 2A**). BsAb was associated with rapid CD4+ and CD8+ T cell activation, as evidenced by an increase in expression of activation markers (PD-1, ICOS, CD39) within 2 days of treatment initiation (**Figure 2A and Supplementary Figure 2B,C**), as well as both a decrease in the naïve T cell fraction and an increase in the proliferative index, which was more pronounced in CD8+ T cells (**Figure 2B**). Interestingly, all T cell subsets other than naïve, central-memory and regulatory T cells showed an increase in Ki-67 positivity at day 2 post-BsAb treatment (**Figure 2C**), but only effector-memory and cytotoxic T cells significantly increased in abundance following BsAb treatment. This suggested that intratumoral accumulation of T cell subsets following BsAb therapy requires both activation and sufficient T cell recruitment (**Supplementary Figure 2D,E**). Analysis of changes in T cell subtype abundance across spleens, LN, and bone marrow revealed a consistent decrease in naïve T cells but tissue-specific changes in other tissues. Central memory T cells increased specifically in the LN, while no substantial changes in effector or memory subsets were observed in the BM, suggesting tissue-specific accumulation of T cell subsets following BsAb treatment (**Figure 2D,E**).

**Figure 2.**
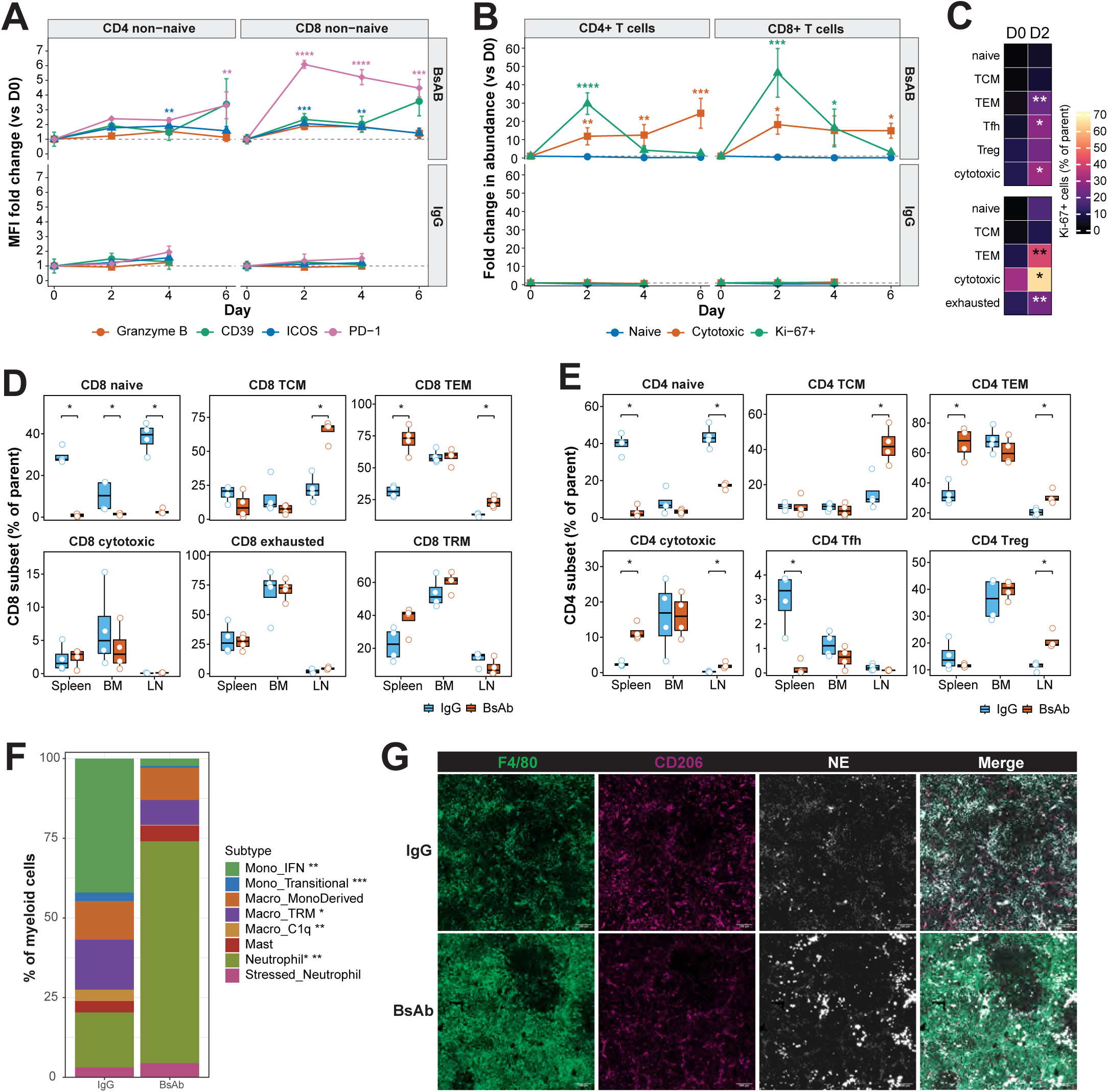
BsAb treatment induces broad T-cell activation and time-dependent myeloid remodeling. (A) Fold change relative to day 0 in the abundance of naive, cytotoxic, and Ki-67⁺ CD4⁺ and CD8⁺ T-cell populations following IgG control or BsAb treatment. (B) Fold change relative to day 0 in the mean fluorescence intensity (MFI) of granzyme B (GranB), CD39, ICOS, and PD-1 in non-naïve CD4⁺ and CD8⁺ T cells. For (A–B), data are shown as mean ± SEM (n =3, day 0; n =5, day 2 IgG; n =6, day 4 after IgG; n =4, day 2 BsAb; n =6, day 4 BsAb; n =3, day 6 after BsAb treatment). No day 6 IgG samples were available because of mortality. Significance was assessed by linear modeling of log₂-transformed abundance (A) or MFI (B), with planned comparisons versus day 0 and Dunnett adjustment for multiple comparisons. *p< 0.05, **p< 0.01, ***p< 0.001, ****p< 0.0001. (C) Percentage of Ki-67+ cells within the indicated CD4 and CD8 T-cell subsets at baseline and 2 days after BsAb treatment (n=3 day 0; n=4 day 2 BsAb). P values were determined using two-sided unpaired t-tests with Benjamini–Hochberg correction for multiple comparisons; *p<0.05, **p<0.01. (D,E) CD8 (D) and CD4 (E) T-cell subset composition in the spleen, bone marrow, and lymph nodes of IgG- and BsAb-treated mice on day 4 (n =4 per group). Each subset is shown as a percentage of its parent population. Boxes indicate the median and interquartile range, and points represent individual mice. P values calculated using two-sided Wilcoxon rank-sum tests with Benjamini–Hochberg adjustment for multiple comparisons. *p< 0.05, **p< 0.01, ***p< 0.001. (F) Myeloid subtype composition by treatment condition (IgG vs. BsAb) in tumor-bearing mice after 4 days of treatment (n = 4 mice/group), determined by single-cell RNA/CITE sequencing with subtypes annotated as in Supplementary Figure 2I, J. Asterisks indicate differential abundance (BsAb vs. IgG) by propeller (moderated t-test on arcsine square-root-transformed proportions): *p< 0.05, **p< 0.01, ***p< 0.001. (G) Representative multiplexed immunofluorescence images of additional sections from spleens of tumor-bearing mice, harvested 4 days after IgG or BsAb treatment, showing F4/80 (green), CD206 (magenta), neutrophil elastase (NE; white), and composite overlays. Scale bars, 100 μm.

To further characterize BsAb-induced T-cell remodeling, CD8+ and CD4+ T-cell subsets were assessed across tissues on day 4. BsAb treatment reduced naive CD8+ T cells across all three tissues (spleen, 27.8% to 0.93%; bone marrow, 10.3% to 1.53%; lymph nodes, 39.6% to 2.13%), accompanied by increased CD8+ TEM cells in the spleen (31.4% to 73.3%) and LN (13.5% to 22.7%) and increased CD8⁺ TCM cells in the LN (21.0% to 68.2%; all p<0.05; **Figure 2D**). A similar shift was observed among CD4+ T cells, with reduced non-Treg naive cells and increased TEM and cytotoxic populations in the spleen and lymph nodes. BsAb treatment also increased TCM and Treg cells in the lymph nodes while reducing splenic Tfh cells (p=0.021; **Figure 2E**). Together, these findings demonstrate broad, tissue-dependent remodeling of CD4 and CD8 T-cell compartments toward activated and memory states following BsAb treatment.

Our immune profiling experiments suggested that BsAb therapy increased not only T cell abundance, but also myeloid cell abundance across tissue types that was most prominent at 6 days following treatment (**Figure 1I and Supplementary Figure 1G,H**). To better characterize changes in myeloid populations with BsAb therapy, we applied an optimized 16-marker myeloid flow cytometry panel to identify monocyte, granulocyte and dendritic cell subsets (**Supplementary Figure 2F,G and Supplementary Table 1**). Myeloid profiling demonstrated a modest increase across myeloid subsets, the most prominent of which were classical monocytes and Ly6G+ PMNs at predominantly 6 days following BsAb treatment (**Supplementary Figure 2H**). To determine whether BsAb promotes phenotypic differences in the myeloid cell compartment, we analyzed CITE-seq data of myeloid cells isolated from tumor-bearing mice at D4 after IgG or BsAb treatment (n=5,913 cells) (**Supplementary Figure 2I**). Unsupervised clustering resolved eight myeloid subtypes, including inflammatory (Mono_IFN) and transitional monocytes, three macrophage populations (Macro_C1q, Macro_TRM, and Macro_MonoDerived), mast cells, and two neutrophil populations (Ly6G+ Neutrophil and Stressed_Neutrophil) (**Supplementary Figure 2I-J**). BsAb treatment was associated with a substantial reduction in inflammatory monocytes (37.5% to 2.3%; propeller, P = 2.2×10⁻³) with a reciprocal expansion of neutrophils (19.5% to 70.0%, propeller p = 1.7×10⁻⁴) (**Figure 2F**). Gene set enrichment analysis of myeloid cells confirmed a significant decrease in interferon signaling across myeloid subclusters (**Supplementary Figure 2K**). Targeted gene set enrichment analysis demonstrated a significant enrichment in genes related to Fc receptor engagement, opsonization, and phagocytosis, suggesting that neutrophils contribute to clearance of BsAb-bound apoptotic tumor cells (**Supplementary Figure 2L**). Consistent with this hypothesis, multiplexed immunofluorescence of spleens from tumor-bearing mice treated with control antibodies or BsAb using myeloid-focused panel including F4/80, CD206, and neutrophil elastase showed a specific increase in neutrophil elastase-expressing granulocytes following BsAb therapy (**Figure 2G**).

### CD3xCD20 BsAb efficacy requires T cell engagement by CD8⁺ but not CD4^+^ T cells *in vivo*

CD3xCD20 BsAb are distinguished from conventional CD20 antibodies by the presence of a CD3 agonist arm. However, both BsAb and conventional anti-CD20 antibodies can engage the immune system through Fc receptor engagement. To distinguish the immunological effects of BsAb and CD20 engagement alone, we compared tumor-bearing mice treated with BsAb to mice treated with a murine anti-CD20 antibody that, unlike BsAb, retains Fc receptor binding capacity due to the lack of LALAPG modification ^32^ (**Figure 3A**). Treatment with this antibody significantly reduced the burden of malignant B cells in the spleen but did not substantially prolong survival compared with IgG control treatment (p=0.464) (**Supplementary Figure 3A,B**). Additionally, only BsAb but not anti-CD20 treatment induced T cell activation and effector differentiation as evidenced by increased expression of CD44, CXCR6, ICOS, PD-1, CD39, and CX3CR1 in BsAb treated but not anti-CD20 treated mice (**Figure 3B and Supplementary Figure 3C**)

**Figure 3.**
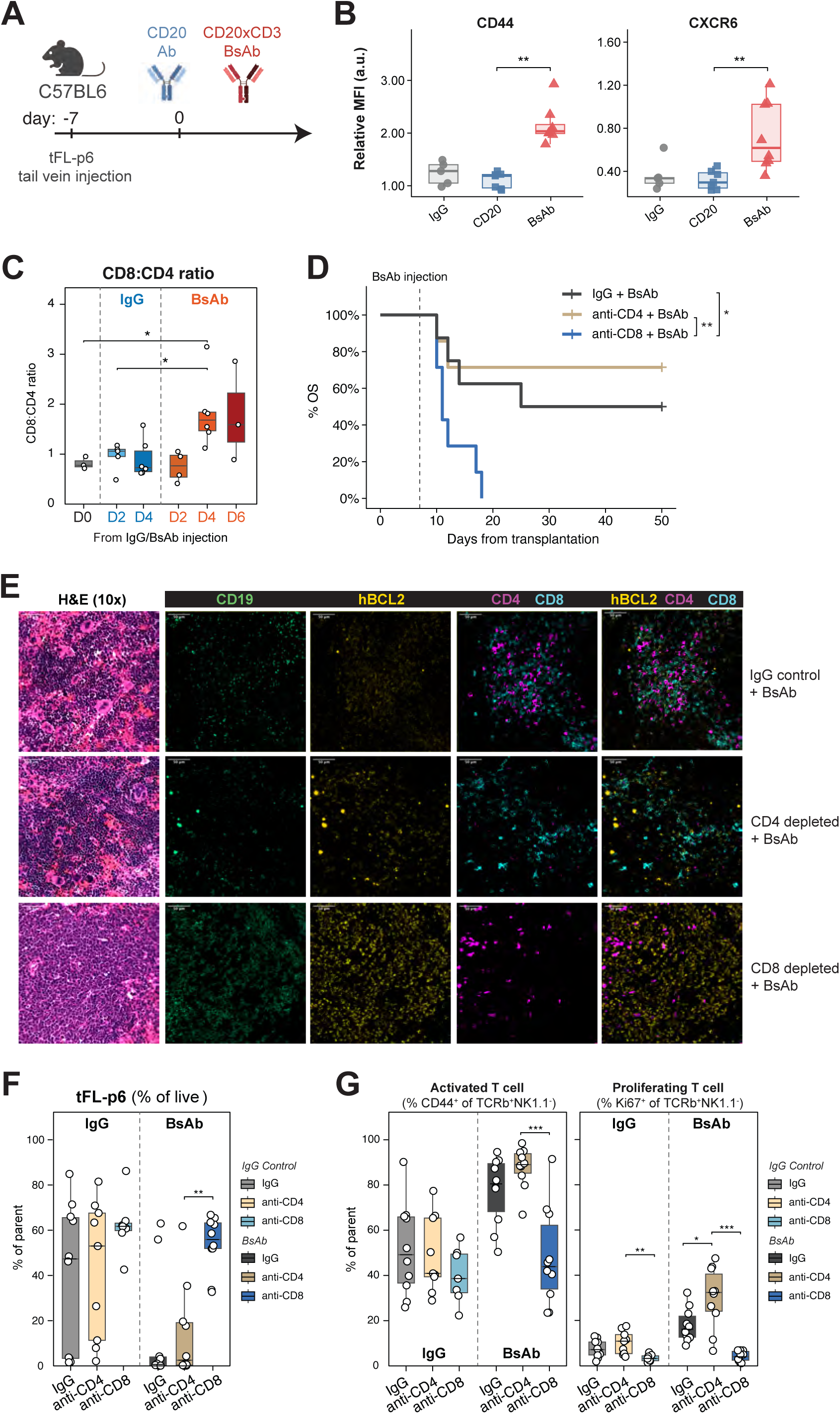
BsAb-specific T-cell activation and the essential role of CD8⁺ T cells for BsAb efficacy *in vivo*. (A) Experimental schema. (B) Relative mean fluorescence intensity (MFI) of CD44 and CXCR6 on splenic NK1.1⁻TCRβ⁺ T cells from IgG-, anti-CD20-, and BsAb-treated mice (n=5, 7, and 8, respectively). Median and interquartile range are shown, points represent individual mice. P values were calculated using two-sided Wilcoxon rank-sum tests comparing anti-CD20- and BsAb-treated mice, with Benjamini–Hochberg correction. **p<0.001. (C) Splenic CD8:CD4 ratio over time following IgG control or BsAb treatment (day 0, day 2 IgG, day 2 BsAb, day 4 IgG, day 4 BsAb, day 6 BsAb). Each point represents an individual mouse; boxplots show median and IQR. Differences across conditions were significant by Kruskal–Wallis test (p=0.023); p values determined by Wilcoxon rank-sum test; *p<0.05. (D) Survival of mice treated with BsAb (left) or IgG control (right) in the presence of CD4 depletion, CD8 depletion, or isotype control. N = 7–8 per group (IgG+BsAb, n=8; CD4-depleted+BsAb, n=7; CD8-depleted+BsAb, n=7; IgG+IgG, n=7; CD4-depleted+IgG, n=8; CD8-depleted+IgG, n=7). Log-rank test; *p<0.05; **p<0.01. (E) Representative multiplexed immunofluorescence of spleens harvested 4 days after BsAb treatment with concurrent IgG control, CD4 depletion, or CD8 depletion, showing H&E (10x) and CD19, hBCL2, CD4, and CD8 staining with composite overlays. (F–G) Spectral flow cytometry-based quantification of splenic tumor burden (tFL-p6; %GFP of live) (E) activated T cells (CD44+), and proliferating T cells (Ki-67+) (F) in mice treated with BsAb or IgG control with or without CD4 or CD8 depletion at day 4 after BsAb treatment (IgG+IgG, n=10; CD4-depleted+IgG, n=9; CD8-depleted+IgG, n=7; IgG+BsAb, n=10; CD4-depleted+BsAb, n=10; CD8-depleted+BsAb, n=10). Each point represents an individual mouse. P values calculated by Wilcoxon rank-sum test; *p<0.05; **p< 0.01, ***p <0.001.

Because BsAb treatment preferentially expanded and activated CD8+ T cells, we next quantified the intratumoral balance of CD8+ and CD4+ T cells following BsAb treatment. We found that the CD8:CD4 ratio progressively increased following BsAb but not control IgG treatment (day 4: 1.82 vs. 0.91, p = 0.026) (**Figure 3C**). To test whether BsAb-driven CD8+ T cell activation is required for therapeutic efficacy, we treated tFLp6-bearing mice with depleting antibodies against CD8 or CD4 prior to BsAb or IgG control treatment (**Supplementary Figure 3D,E**). Survival analysis showed that CD8 depletion eliminated the therapeutic benefit of BsAb (median OS 11 vs. 25 days with BsAb + anti-CD8 vs. BsAb + IgG, p=0.003) (**Figure 3D**). In contrast, BsAb remained effective in CD4-depleted mice (median OS not reached with BsAb + anti-CD4 vs. 25 days with BsAb + IgG, p = 0.496) (**Supplementary Figure 3F**).

To determine how loss of CD8+ T cells abrogated BsAb efficacy, we analyzed the tumor microenvironment of Bcl2/Ezh2 tumors following either BsAb or isotype control treatment. In T cell replete mice, BsAb treatment was associated with reduced hBCL2/CD19 tumor signal and prominent accumulation of primarily CD8+ T cells in and around tumor regions (**Figure 3E**). CD4 depletion had minimal impact on this spatial pattern, whereas CD8 depletion clearly diminished intratumoral T cell presence and was accompanied by persistence of hBCL2/CD19 tumor areas, suggesting that BsAb-mediated tumor control in this model is primarily CD8 T cell–dependent. This was specific to BsAb treatment, as CD8 depletion did not alter the tumor spatial pattern in mice treated with isotype control antibodies (**Supplementary Figure 3G**). Single-cell immune profiling of spleens 4 days after treatment confirmed that CD8+ T cell depletion completely abrogated the reduction in tumor burden induced by BsAb therapy (**Figure 3F**). In CD4-depleted mice, BsAb-driven T cell activation (CD4-depleted vs IgG control: 87.7% vs 77.5%, p=0.968) and proliferation (30.1% vs 17.9%, p=0.243) were similar to non-T cell depleted mice, and T cell phenotypes were largely unaffected (**Figure 3G and Supplementary Figure 3H,I**). In contrast, CD8 depletion prior to BsAb therapy significantly reduced T cell activation (48.2% vs 87.7%, p<0.0001) and proliferation (4.2% vs 30.1%, p<0.0001) compared with CD4 depletion (**Figure 3G and Supplementary Figure 3H,J**).

### Clonally expanded cytotoxic CD8+ T cells accumulate in patients with durable response to CD3xCD20 BsAb therapy

Given the dependence of BsAb efficacy on CD8+ T cells in Bcl2/Ezh2 tumor-bearing mice, we asked whether intratumoral T cell composition was associated with clinical response in patients treated with CD3xCD20 bispecific antibodies. We began by analyzing clinical flow cytometry data from patients with FL who received epcoritamab-based combination therapy as part of a multi-arm clinical trial (NCT #04663347, Arms 2 and 6, Rituximab + Lenalidomide + Epcoritamab^33^) (**Figure 4A; Supplementary Table 2**). Compared with patients who remained progression-free, pre-treatment samples from 2 patients who experienced PD (one of whom had previously achieved CR) had substantially fewer intratumoral CD8^+^ T cells and a lower CD8:CD4 T cell ratio (**Figure 4B** and **Supplementary Figure 4A**). This difference persisted following treatment, suggesting that increased intratumoral CD8 T cell abundance may enable durable remissions after epcoritamab-based therapies.

**Figure 4.**
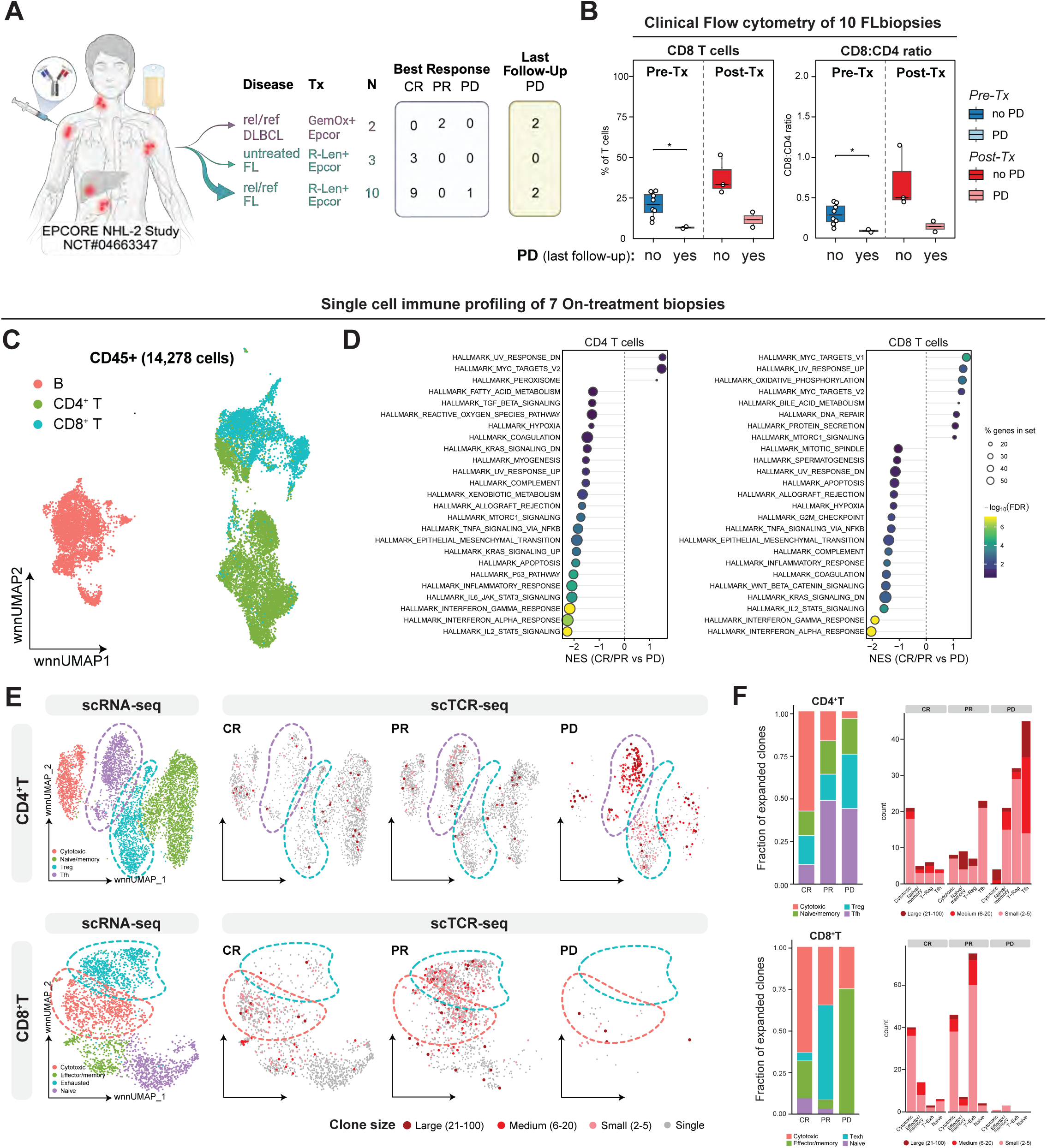
CD8 and cytotoxic T cell expansion in patients with durable responses to BsAb. (A) Overview of the clinical cohorts and sampling strategy from the EPCORE NHL-2 study (NCT04663347), including disease setting, treatment regimen, number of patients, and best response categories (CR, PR, PD). (B) Intratumoral T cell composition before and after BsAb-based therapy, stratified by clinical response. Boxplots show the fraction of CD8 T cells (left) and CD8:CD4 ratio (right) in samples from responders (no PD) versus the single patient with PD. Each point represents one sample/patient. P values calculated using Wilcoxon rank-sum test; *p < 0.05. (C) wnnUMAP of tumor-infiltrating CD45+ immune cells colored by major lineages (B cells, CD4 T cells, CD8 T cells). (D) Gene set enrichment analysis (Hallmark pathways) comparing responders (CR/PR) versus PD within intratumoral CD4 T cells (left) and CD8 T cells (right). Dots denote enriched pathways; x-axis shows normalized enrichment score (NES), dot size indicates the fraction of genes in the set contributing to the signal, and color denotes significance (−log10 FDR). (E) Joint single-cell transcriptome and TCR profiling of intratumoral CD4 T cells (top) and CD8 T cells (bottom). For each compartment, wnnUMAPs show transcriptional states (left) and expanded clonotypes overlaid for representative CR, PR, and PD cases (right; expanded clones highlighted). Clone size categories are indicated in the legend. (F) Quantification of clonal expansion by response category. Stacked bars show the fraction of expanded clonotypes distributed across cell states in CD4 T cells (top) and CD8 T cells (bottom) (left panels). Right panels show the distribution of clone sizes per response group.

Next, to determine whether response to epcoritamab-based therapies was associated with quantitative or qualitative differences in intratumoral T cell phenotypes, we analyzed dissociated single cells from on-treatment biopsies collected from 7 patients with available material (2 with previously untreated FL, 3 with R/R FL - all 5 receiving a combination of epcoritamab with lenalidomide and rituximab - and 2 with R/R DLBCL having received epcoritamab, gemcitabine and oxaliplatin^36^) using the 10x Genomics Single Cell Immune

Profiling platform (**Supplementary Table 2**). After quality control filtering, we analyzed a total of 14,278 cells from these 7 patients (**Figure 4C**). Seurat-based clustering of CD4 and CD8 T cells identified known lymph node-resident phenotypes including naïve CD4 and CD8 T cells (expressing *IL7R, SELL, CCR7*), cytotoxic CD4 T cells (*GZMK, PRF1, NKG7*), follicular helper CD4 T cells (Tfh) (*CXCL13, TOX2, PDCD1*), regulatory T cells (Treg) (*FoxP3, Il2RA, TNFRSF1B*), cytotoxic CD8 T cells (*S100A4, CXCR6, PRF1*), effector/memory CD8 cells (*IL7R, SELL, S100A4, CX3CR1*), and exhausted CD8+ T cells (*PDCD1, TOX, CXCL13*) (**Supplementary Figure 4B-D**). Hallmark gene set enrichment analysis (GSEA) of CD4+ and CD8+ T cells from patients with complete or partial response (CR/PR) versus progressive disease (PD) revealed enrichment of gene sets associated with metabolic fitness (HALLMARK_MYC_TARGETS, HALLMARK_OXIDATIVE_PHOSPHORYLATION) in durable responders and chronic inflammation (INFLAMMATORY_RESPONSE, IL6_JAK_STAT3_SIGNALING, INTERFERON_GAMMA_RESPONSE, INTERFERON_ALPHA_RESPONSE, IL2_STAT5_SIGNALING) in the single PD biopsy (**Figure 4D, Supplementary Figure 4E**). These data suggest that durable responses are linked to a metabolically fit, proliferative T cell state, whereas treatment failure is characterized by a chronically inflamed, interferon-dominated transcriptional program in both CD4+ and CD8+ T cells.

Finally, to determine whether response to BsAb was associated with differences in clonal expansion of intratumoral T cells, we analyzed combined CITE-seq and scTCR sequencing data from on-treatment biopsies. This revealed cell state-specific clonal expansion in both the CD4+ and CD8+ T cell compartments (**Figure 4E,F**). In patients achieving a durable CR, most clonally expanded T cells in both the CD4+ and CD8+ T cell compartments showed a cytotoxic phenotype, with expression of *CST7, CCL4,* and *NKG7* (**Figure 4E** and **Supplementary Figure 4F**). In contrast, the patient with PD showed substantial expansion of CD4+ Treg subsets (as has been observed in non-responders to blinatumomab in patients with B-ALL ^37^) but also of Tfh cells with almost no CD8+ T cell expansion. Patients achieving a PR exhibited an intermediate phenotype with less expansion of cytotoxic T cells compared to CR patients and clonal expansion of CD8+ T cells with an exhausted phenotype. Analysis of clonal expansion across patients and cell states showed that lasting CR was associated with clonal expansion of cytotoxic CD4+ and CD8+ T cells. PR showed an intermediate pattern, with less cytotoxic expansion than CR patients but greater clonal expansion of exhausted CD8+ T cells along with highly expanded non-cytotoxic CD4+ T cell clones^38, 39^. The single PD case showed minimal CD8 expansion but marked expansion of large Tfh clones (**Figure 4E,F**). Taken together, these results indicate that a durable response to BsAb in patients with B cell lymphomas is associated with an increase in polyclonally expanded intratumoral cytotoxic CD8+ T cells.

### CXCR6-dependent T cell recruitment enables optimal CD3xCD20 BsAb efficacy in vivo

In tFLp6-bearing mice, BsAb therapy promoted intratumoral accumulation of cytotoxic CD8+ as well as CD4+ T cells. We observed similar findings in patients with prolonged CR after BsAb-based therapy, in whom cytotoxic CD4+ and CD8+ T cells showed substantial intratumoral clonal expansion. To directly validate the role of distinct T cell subsets in mediating BsAb activity, we performed *in vitro* cytotoxicity assays using polarized human T cell subsets. CD4+ and CD8+ T cells were activated under polarizing conditions for 5 days ^40, 41^, sorted, and subsequently co-cultured with luciferase-expressing RAJI B cells in the presence or absence of the BsAb epcoritamab (**Supplementary Figure 5A,B**). We observed no significant B cell killing by any polarized T cell subset in the absence of epcoritamab (**Supplementary Figure 5C**). BsAb addition induced significant tumor cell killing by both CD8+ and Th1-polarized CD4+ T cells, while CD4 Tregs and Tfh cells exhibited minimal or no cytotoxic activity (**Figure 5A**).

**Figure 5.**
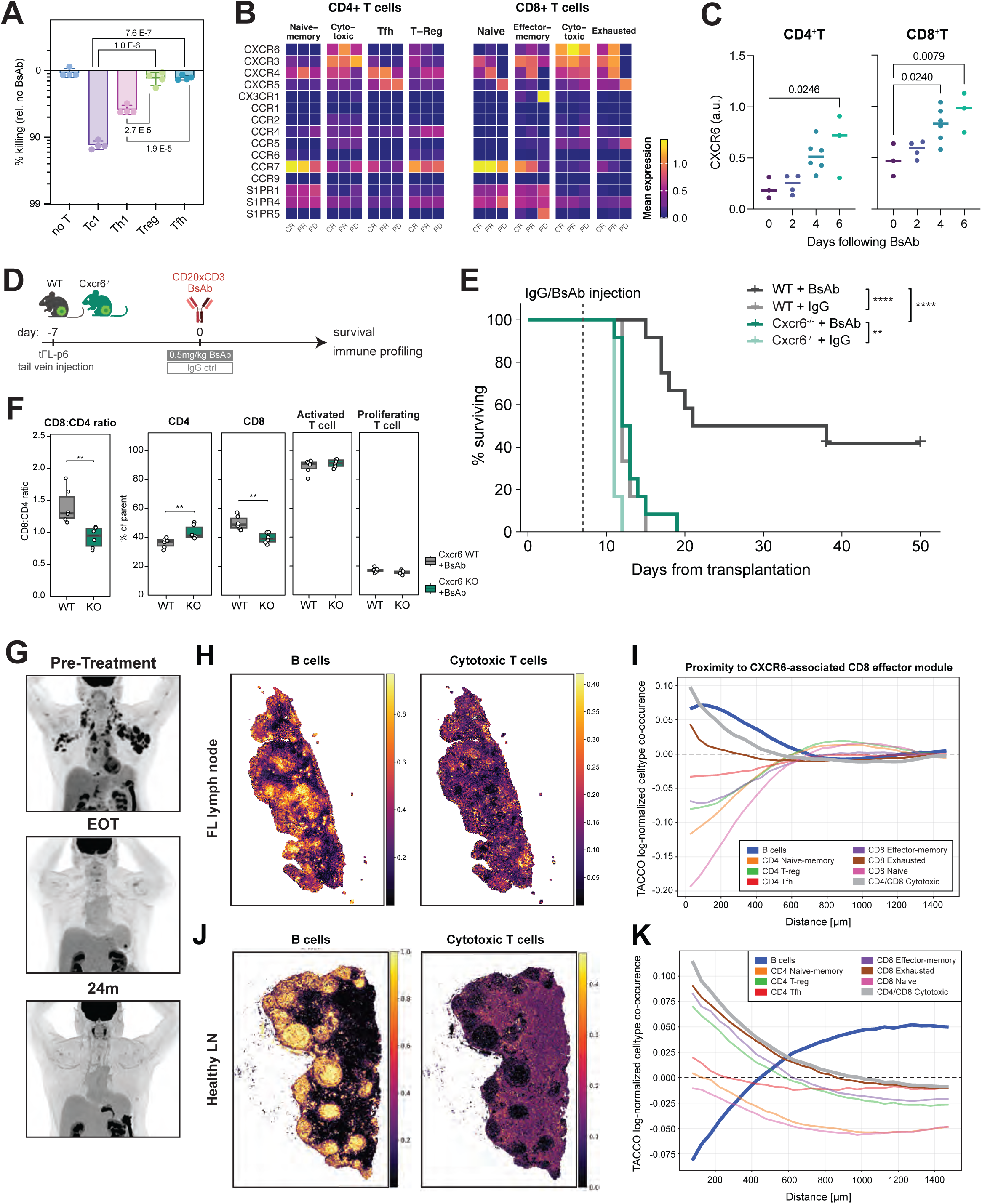
CXCR6 is essential for BsAb-driven tumor infiltration and immune-mediated tumor control. (A) Quantification of tumor cell killing by in vitro–polarized human T cell subsets co-cultured with luciferase-expressing Raji B cells ± epcoritamab. P values determined by one-way ANOVA with Sidak correction for multiple comparisons. (B) Chemokine receptor expression across CD4⁺ and CD8⁺ T-cell subtypes stratified by clinical response. Heatmaps show mean normalized single-cell RNA expression of 15 chemokine and sphingosine-1-phosphate receptors in intratumoral CD4⁺ T-cell subtypes and CD8⁺ T-cell subtypes, grouped by clinical response (CR, PR, or PD). Color scale indicates mean expression within each subtype-response group. (C) Expression levels of CXCR6 by spectral cytometry in splenic CD4+ and CD8+ T cells at the indicated timepoints following BsAb therapy. P values determined by one-way ANOVA with Sidak correction for multiple comparisons. (D) Experimental design for testing CXCR6 function in BsAb response. (E) Kaplan–Meier survival of tumor-bearing WT and Cxcr6 knockout mice treated with BsAb or IgG control (n = 6/group). P values calculated by Log-rank test; *p < 0.05, **p < 0.01, ***p < 0.001, ****p < 0.0001. (F) Spectral flow cytometry of splenocytes after BsAb treatment in WT versus Cxcr6 knockout mice (n = 6/group). Left, CD8:CD4 ratio. Right, Frequencies of CD4, CD8, activated T cells and proliferating T cells, defined by CD44 and Ki-67 expression, respectively, within the T cell compartment. P values calculated by Wilcoxon rank-sum test; **p<0.01. (G) PET-CT scans for a patient achieving a prolonged CR following epcoritamab, rituximab, and lenalidomide at indicated timepoints. (H) Spatial transcriptomic profiling of an on-treatment right cervical lymph node biopsy from a patient with follicular lymphoma (FL) who achieved a prolonged complete response following epcoritamab, rituximab, and lenalidomide. Spatial distribution of B cell (left) and cytotoxic T cell (right) programs from Figure 4E and **Supplementary Figure 4B-C**. (I) Spatial proximity analysis of the FL lymph node, showing the proximity of inferred cell types to the CXCR6-associated CD8 effector module. (J) Spatial transcriptomic profiling of a healthy (non-malignant) lymph node, showing the spatial distribution of B-cell (left) and cytotoxic T-cell (right) programs. (K) Spatial proximity analysis of the healthy lymph node, showing the proximity of inferred cell types to the CXCR6-associated CD8 effector module.

To identify chemokine receptors associated with cytotoxic and effector T-cell states, we systematically compared the expression of 15 chemokine receptors (CXCR3–6, CX3CR1, CCR1/2/4/5/6/7/9, and S1PR1/4/5) across CD4+ and CD8+ T-cell subsets in human scRNA/CITE-seq data (**Figure 5B**). CXCR6, a chemokine receptor implicated in T-cell trafficking to inflamed tissues and tumors, emerged as the receptor most consistently enriched in cytotoxic/effector populations across both lineages. Consistent with this finding, *CXCR6* expression was strongly induced in both CD4+ and CD8+ T cells beginning at D4 after BsAb treatment in tumor-bearing mice (**Figure 5C**).

Given these results, as well as reports linking CXCR6 expression to increased intratumoral localization of cytotoxic T cells as well as CAR-T cells ^42, 43, 44, 45^, we hypothesized that CXCR6 may be functionally required for an effective CD8+ T cell response to BsAb therapy *in vivo.* We therefore challenged wild-type and Cxcr6^-/-^ mice (JAX #005693) with systemic Bcl2/Ezh2 lymphoma (tFLp6; day −7 tail-vein injection) and treated with BsAb or IgG control (**Figure 5D**). BsAb treatment of tumor-bearing Cxcr6-deficient mice showed a small though statistically detectable survival benefit (median OS 12.5 days vs. 11 days with IgG control treatment; p<0.01), that was markedly inferior to the benefit in wild-type mice (median OS 12.5 vs. 21 days; p<0.0001) (**Figure 5E**). Single-cell immune profiling of spleens by spectral flow cytometry showed that, following BsAb treatment, Cxcr6^-/-^ mice had a significantly reduced intratumoral CD8:CD4 ratio compared with WT ones, despite no detectable differences in the overall frequency of activated or proliferating T cells (**Figure 5F**) and modest differences in expression of other phenotypic markers (**Supplementary Figure 5D**), suggesting that CXCR6 may be dispensable for BsAb-driven T cell activation but is important for intratumoral accumulation and/or localization. The difference in intratumoral accumulation in CD8+ T cells was specific to BsAb therapy, as untreated tumor-bearing WT and Cxcr6^−/−^ mice showed broadly comparable CD4+ and CD8+ T-cell subset distributions with the exception of known CXCR6+ populations (cytotoxic CD4s, Cytotoxic CD8s, CD8 TRM) (**Supplementary Figure 5E**).

The dependence of BsAb efficacy on CXCR6 expression indicated that BsAb efficacy requires not only activation but recruitment and apposition of cytotoxic T cells to malignant B cells within the tumor microenvironment. To test this hypothesis, we performed spatial single-cell transcriptomic profiling using the 10x Genomics Visium HD platform on a biopsy obtained from a patient with follicular lymphoma following initiation of treatment with rituximab, lenalidomide, and epcoritamab who had pathologically documented residual malignant B cells at 3 weeks following treatment but went on to achieve a complete metabolic response and has remained free of relapse nearly 3 years on (**Figure 5G and Supplementary Figure 5F**). We applied the Transfer of Annotations to Cells and their Combinations (TACCO) framework that annotates expressed genes within spatial transcriptomic spots by their probability of belonging to any of the 9 refined cell type annotations from our single cell immune profiling dataset and observed overlapping pockets of predictions for malignant B cells and cytotoxic T cells (**Figure 5H**). Next, we derived a *CXCR6-associated CD8 cytotoxic gene module* based on differentially expressed genes between CXCR6-high and CXCR6-low CD8+ T cells to assess its spatial distribution within the biopsied lymph node **(Supplemental Figure 5G)**. We then quantified spatial proximity between module-high spots and inferred cell types by measuring distances to the nearest B cell and cytotoxic T cell regions. Interestingly, aside from cytotoxic T cells themselves, module-high regions were closest to B cell regions, consistent with BsAb treatment promoting close apposition of malignant B cells and CXCR6/cytotoxic T cells **(Figure 5I**).

To confirm that this co-localization was specific to BsAb treatment, we applied a similar analysis to publicly available spatial transcriptomic data from a healthy human lymph node sample (**Figure 5J**). In contrast to the sample after BsAb treatment, B cell predictions minimally overlapped with those for cytotoxic T cells (**Figure 5J**) and, correspondingly, showed the least proximity to the CXCR6/cytotoxic module (**Figure 5K**). These data underline that optimal response to BsAb therapy likely requires CXCR6-dependent recruitment of cytotoxic T cells to tumor-rich regions.

## DISCUSSION

Bispecific T cell engagers represent a novel, highly effective therapeutic strategy in patients with B cell lymphoma, with encouraging single-agent clinical results in the relapsed/refractory setting ^14, 15^. They provide an off-the-shelf immunotherapeutic strategy for patients with lymphoma, which is particularly valuable given that immune checkpoint blockade with antibodies targeting either PD-1 or CTLA-4 have shown minimal activity in non-Hodgkin B cell lymphomas. Understanding determinants of response and resistance is, therefore, crucial for the continued development of these agents. Interestingly, while antigen loss as a mechanism of disease progression has been described, many patients’ tumors retain antigen expression at the time of disease progression ^46^, suggesting alternative drivers of BsAb resistance. We hypothesized that intratumoral immune cell composition would influence BsAb efficacy, given that it is known to impact lymphoma development and progression ^47^, and that BsAb non-specifically engage any cell expressing CD3 epsilon.

Our study has several major findings. First, we establish an immunocompetent BCL2/Ezh2-driven syngeneic lymphoma model in which CD3xCD20 BsAb therapy induced rapid tumor clearance. This model has the advantage of being syngeneic with the C57/Bl6 mice, enabling it to be combined with nearly all mouse strains with genetically inbred immunological defects to validate immune drivers of BsAb sensitivity. The sensitivity of the model to BsAb therapy makes this platform highly amenable to genetic screens for tumor cell-autonomous drivers of BsAb resistance.

Second, we showed that BsAb efficacy depends on engagement of T cells, and specifically CD8+ T cells, while CD4+ T cells were dispensable for tumor elimination. Our observation of intratumoral accumulation of BsAb-unresponsive CD4+ cell states, including Tfh and Treg, in the single patient who was refractory to BsAb may be particularly relevant in FL, where Tfh programs have been shown to support disease progression ^20, 48, 49, 50, 51, 52^.

Whether Tfh persistence or proliferation following BsAb therapy might enable lymphoma progression, as suggested by a recent report ^53^, is an intriguing topic for future study.

Regardless, our results suggest that strategies to limit engagement of non-cytotoxic CD4+ T cells by BsAb, for example via Notch inhibition to limit Tfh differentiation^54^, may enhance response to treatment. For example, Notch inhibition may promote differentiation of CXCR6+ CD8+ T cells while limiting accumulation of CD4+ Tfh cells^54, 55^.

Third, we identify CXCR6 expression as necessary for effective BsAb responses in lymphoma tumor-bearing mice and show preferential CXCR6+ clonal T cell expansion in patients with complete remissions to BsAb-based therapies, suggesting that durable BsAb responses require ongoing chemokine-dependent recruitment of cytotoxic T cells to tumor-proximal regions. CXCR6-mediated T cell recruitment has been implicated in anti-tumor immunity and response to immunotherapy in solid tumors^56^ as well as in the context of cellular immunotherapies ^44, 45^. Immune checkpoint blockade is increasingly appreciated to act on progenitor-like exhausted T cells within tumor-draining lymph nodes, driving LN egress and tumor infiltration^57, 58^; as such it is logical that efficacy would depend on chemokine receptor-mediated recruitment. In contrast, CD3xCD20 BsAb putatively supply both target specificity and activating signals to any CD3 epsilon-expressing T cell. In this context, it is surprising that BsAb efficacy was almost completely dependent on CXCR6-mediated T cell recruitment. This represents a novel constraint specific to the pharmacology of T cell engagers in which their efficacy is limited not purely by T cell activation but by whether activated cytotoxic T cells reach and are retained at tumor-proximal sites. Whether strategies to increase CXCR6-CXCL16 driven T cell recruitment may also enhance BsAb responses remains to be investigated. However, this should be approached carefully given emerging evidence linking CXCR6-CXCL16 activation to neurotoxicity in the context of cellular therapies^59^.

We acknowledge the limitations of this study, including the modest sample size of the patient cohort, as well as the fact that epcoritamab was administered in combination with other systemic agents in the patients studied. In addition, the mouse experiments lacked binding-arm-matched controls, limiting our ability to fully exclude effects attributable to either targeting arm independently of simultaneous CD3 and CD20 engagement.

Nonetheless, by integrating causal depletion experiments in an immunocompetent lymphoma model with single-cell and TCR-resolved profiling of human tissue, our study provides a mechanistic rationale for therapeutic strategies that preferentially recruit and sustain cytotoxic CD8+ T-cell programs, including tissue-associated CXCR6+ states, while minimizing nonproductive BsAb engagement of CD4+ T-cell states.

## METHODS

### Patients

NCT #04663347 is a multi-arm phase 1/2 international trial of epcoritamab in combination with various standard-of-care therapies for patients with DLBCL or FL. Patients included in the present analysis included patients with DLBCL, who received epcoritamab in combination with either R-CHOP (newly diagnosed) or gemcitabine and oxaliplatin (relapsed/refractory), or FL, who received epcoritamab in combination with rituximab and lenalidomide (**Figure 4A and Supplementary Table 2**). Eligibility criteria, treatment schedule, and preliminary safety and efficacy results for each of these study arms were previously reported^33, 34, 60, 61^. As part of the study procedures, several patients underwent optional on-treatment biopsies during cycle 1 between days 22 and 28. Some subjects also underwent biopsy at the time of progression. Each patient provided written consent to participate in the study and had voluntarily opted into additional research biopsies. The study was conducted in accordance with the principles outlined in the Declaration of Helsinki and was approved by the MSKCC Institutional Review Board (IRB #21-164).

### Multiparametric Flow Cytometry

Clinical flow cytometry was performed as previously described ^62^. Briefly, tissue was finely minced in RPMI 1640 growth medium using scalpel blades, then filtered, washed, and resuspended in RPMI. Following resuspension, cells were stained with a 21-antibody panel designed to assess B- and T cell subsets (**Supplementary Table 2**) and incubated at room temperature. Following staining and incubation, cells were lysed, fixed, washed, and resuspended. Up to 500,000 cells were acquired on a FACSymphony A3 flow cytometer (BD Biosciences). The results were analyzed with Woodlist software (Dr. B.L. Wood, University of Southern California).

### Immunophenotyping

Cells were labeled with viability dye (Zombie NIR, BioLegend #423105) for 10 min at 4°C and stained with surface antibodies for 30 min at room temperature. Cells were then fixed/permeabilized (Thermo Fisher Fixation/Permeabilization Kit, #00-5523-00) for 30 min at room temperature and stained for intracellular targets overnight at 4°C, followed by fixation in 4% formaldehyde in PBS. Samples were acquired on a Cytek Aurora. Spectral unmixing was performed using single-stained reference controls (and an autofluorescence reference when appropriate), and the resulting unmixing matrix was applied to all samples prior to export of unmixed FCS files for downstream analysis.

High-dimensional cytometry data were analyzed in Omiq (Omiq Inc.). After quality control and hierarchical gating to exclude debris, doublets, and dead cells and to define major immune populations (gating strategy shown in **Supplementary Figure 6**), samples were normalized for visualization by subsampling to a fixed total of 60,000 events across conditions (balanced across conditions/samples as configured in Omiq). The pooled, subsampled dataset was embedded using UMAP on transformed marker intensities, and condition- and timepoint-specific density plots were generated on the shared embedding. Unsupervised clustering was performed in Omiq, and clusters were annotated post hoc based on canonical marker expression using the prespecified gating/marker definitions indicated in the relevant figure and Supplementary methods. Mean fluorescence intensity (MFI) statistics were computed within prespecified gates, and log2 fold-changes were calculated relative to matched IgG controls at each time point per individual experiment.

### Multiplexed immunofluorescence

Paraffin-embedded tissues were sectioned at 5 μm and baked at 58°C for 1 hr. Hematoxylin and eosin (H&E) stains were performed on paraffin-embedded tissue sections following routine processing. Automated multiplexed IF was conducted with the Leica Bond BX staining system. Slides were loaded in Leica Bond and IF staining was performed as follows. Samples were dewaxed at 72 degrees before being pretreated with EDTA-based epitope retrieval ER2 solution (Leica, AR9640) for 20 mins. at 100°C. The 5-plex antibody staining and detection were conducted sequentially. Primary antibodies against Ki67 (Rb, 0.25 μg/mL, Abcam, ab15580), CD4 (Rb, 0.175 μg/mL, Abcam, ab183685), CD8 (Rb, 0.15 μg/mL, Cell Signaling Technology, 98941), BCL2 (Rb, 1:4, Ventana, 790-4604), CD19 (Rb, 0.25 μg/mL, Abcam, ab245235), neutrophil elastase (Rb, 1:500, Abcam, ab68672), F4/80 (Rb, 0.05 μg/mL, Cell Signaling Technology, 70076), CD11b (Rb, 0.125 μg/mL, Abcam, ab133357), CD68 (Rb, 0.5 μg/mL, Boster, PA1518), or CD206 (Rb, 0.125 μg/mL, Abcam, ab64693) were incubated for 1 h at room temperature (RT), followed by application of Leica Bond Polymer anti-rabbit HRP secondary antibody (included in the Polymer Refine Detection Kit; Leica, DS9800) for 8 min at RT. Alexa Fluor tyramide signal amplification reagents (Life Technologies, B40953 and B40958) or CF® dye tyramide conjugates (Biotium, 92172, 96053, and 92174) were then used for detection. After each round of IF staining, epitope retrieval was performed to denature the primary and secondary antibodies before application of the next primary antibody. Following completion of the staining run, slides were washed in PBS and incubated with 5 μg/mL 4′,6-diamidino-2-phenylindole (DAPI; Sigma-Aldrich) in PBS for 5 min, rinsed in PBS, and mounted in Mowiol 4–88 (Calbiochem). Slides were stored overnight at −20 °C before imaging.

### Human Single-cell immune profiling

Single-cell RNA FASTQ data was processed using Cellranger v6.0.2 *count* workflow to generate gene expression count matrices. TCR data was processed using Cellranger’s *vdj* pipeline to generate cell-clonotype annotations.

Processed RNA and ADT matrices were combined into a single Seurat v5.0.1 object (using the *CreateSeuratObject* function), into which TCR clonotype information was subsequently incorporated using *scRepertoire v1.7.2 combineExpression* function. Filtering was applied upon the merged data to only retain cells with (1) greater than 500 and less than 4000 detected unique RNA features, (2) less than 10000 total RNA molecules (3) less than 5 percent mitochondrial RNA reads and (4) less than 2500 total ADT molecules. RNA and ADT data were normalized using Seurat’s *NormalizeData* function with ‘LogNormalize’ and ‘CLR’ methods specified for each, respectively. PCA was performed (using Seurat’s *RunPCA* function) and to mitigate the effect of lane-specific and sample-specific covariates/batch effects; Harmony v1.0.0 *RunHarmony* function was used to correct embeddings. Elbow plots plots were manually inspected to determine the number of principal components to use downstream, and a global UMAP was constructed using Seurat’s weighted nearest neighbor workflow upon corrected RNA and ADT data (n=14,278). Using *clustree v.0.5.0* produced tree diagrams for various resolution input values, cluster stability was evaluated to avoid over-clustering and optimize the number of communities selected. Subsequently, separate Seurat objects for B and T-lymphocytes were produced by evaluation of and isolation by canonical marker expression. T-lymphocytes were further separated by first gating on CD3 and separating into CD8 and CD4 compartments using the ADT expression for each of those markers. Data were filtered to remove cells where ADT and RNA-seq profiles marked different cell types. The previously described UMAP construction and clustering steps were performed for each cell-subtype object. Specialized cell-types were labeled by manually evaluating differentially expressed genes and surface protein markers across clusters. Transcriptional and proteomic differences between treatment and clinical outcome groups were characterized using Seurat’s implementation of the Wilcoxon rank-sum test. Gene set enrichment against hallmark, gene ontology, and canonical pathway terms was computed using the preranked module in gseapy v1.0.6. RPKM-normalized read coverage tracks were generated using pyGenomeTracks v3.8.

### Mouse single-cell immune profiling

Mouse single-cell RNA and ADT data were processed using a workflow analogous to that used for the human dataset, with the following modifications. Reads were aligned using Cell Ranger against a custom reference incorporating the human BCL2 (hBCL2) transgene sequence, allowing malignant tFL-P6 cells to be identified by hBCL2 expression. Count matrices were then imported into Seurat v5.4.0. Cells were retained if they contained >500 and <7,500 detected RNA features and <7.5% mitochondrial reads. Doublets were identified and excluded using scDblFinder v1.24.10, retaining singlets only, and TCR clonotype information was incorporated using the combineExpression function in scRepertoire v2.6.2. RNA and ADT modalities were analyzed independently rather than integrated using a weighted nearest neighbor workflow. Principal component analysis was performed on RNA data alone following ScaleData, with the percentage of mitochondrial reads regressed out, using 50 principal components. No Harmony batch correction was applied because all samples were derived from a single experiment and processed on the same day, thereby minimizing batch-related technical variation. Clustering resolution was selected using a clustree sweep across resolutions 0–1.5 and was set to 1.0. Initial cell-type annotations were generated using SingleR with the ImmGen reference from celldex v1.20.0 and were subsequently manually refined based on differentially expressed genes and surface protein markers, following the same general annotation strategy used for the human dataset.

### Spatial transcriptomic profiling

VisiumHD data sets were processed using SpaceRanger version 3.1.2. Publicly available healthy human lymph node Visium HD data processed by SpaceRanger version 4.0.1 was retrieved from 10X Genomics. All Visium HD data sets were analyzed at 8 μm resolution. We first filtered spots to retain those with mitochondrial reads percentage less than 20% and gene count per cell greater than 50.

The transfer of annotations to cells and their combinations (TACCO) framework ^63^ was subsequently used to compositionally annotate the spots in the Visium HD data sets using the present single cell immune profiling data reported as reference. All cytotoxic CD4/CD8 T cells were relabeled as ‘cytotoxic T cells’. We used the tacco.tools.annotate() function with default parameters and multi_center=3 to compositionally annotate Visium HD spots. Intratumoral CD8+ T cells were classified as CXCR6-expressing or CXCR6-negative based on detectable normalized CXCR6 RNA expression (>0 or 0, respectively). Differential expression was performed using Seurat FindMarkers with a two-sided Wilcoxon rank-sum test, a minimum detection frequency of 10%, and a log2 fold-change threshold of 0.25. P values were adjusted using the Bonferroni method. Based on this analysis, we defined a gene module of CXCR6-associated/effector T cell markers, termed the ‘*CXCR6-associated CD8 effector module’*, and quantified it as the mean expression of *CXCR6, CCL5, CCL4, GZMA, GZMK, NKG7, CST7, IFNG, FASLG, LAG3, PRDM1, CADM1, PLEK, FYN*. We computed the co-occurrence centered on the *CXCR6-associated CD8 effector module* of all reference cell types using the function tacco.tools.co_occurrence() with delta_distance=50 and max_distance=1500.

### *In vivo* experiments

All animal procedures were performed in accordance with institutional guidelines and approved animal care protocols (IACUC protocol 20-10-012, PI: Vardhana). Male mice aged 6-8 weeks old were used for all experiments. The tFL-p6 BCL2/Ezh2 lymphoma cell line was kindly provided by Dr. Beguelin and has been described previously^28, 64^. For syngeneic tumor experiments, C57BL/6J wild-type mice or Cxcr6-deficient mice (B6.129P2-Cxcr6^tm1Litt/J; The Jackson Laboratory, stock #005693) were injected intravenously (tail vein) with 1×10^6^ tFL-p6 BCL2/Ezh2 lymphoma cells on day −7. On day 0, mice received CD20×CD3 bispecific antibody (BsAb; 0.5 mg/kg) or matched IgG control by intravenous tail vein injection and were followed for survival and/or sacrificed at predefined time points for immune profiling. The BsAb and murine anti-CD20 antibody were kindly provided by Genentech ^30^, and comprised a murine IgG2a anti-CD20 arm (clone 5D2) and a hamster anti-CD3 arm (clone 2C11). The BsAb contained LALAPG Fc substitutions to attenuate Fc-mediated effector functions. A matched control antibody was a mouse IgG2a isotype control (InVivoMAb mouse IgG2a isotype control, Bio X Cell, BE0085). Of note, both the IgG2a isotype control antibody and anti-CD20 antibody lack the LALAPG substitutions and are therefore still capable of Fc receptor-dependent immune activation. For time-course immune profiling, spleens were harvested at baseline (day 0) and on days 2, 4, and 6 after treatment (day 6 IgG control not available due to mortality), processed into single-cell suspensions, and analyzed by multiparametric spectral flow cytometry using the antibody panel listed in **Supplementary Table 1**. For T cell depletion studies, mice received an anti-mouse CD4 depleting antibody (InVivoMAb anti-mouse CD4; Bio X Cell, BE0003-1) or an anti-mouse CD8β depleting antibody (InVivoMAb anti-mouse CD8β [Lyt 3.2]; Bio X Cell, BE0223), with matched isotype control, 300 µg i.p. 3 days before BsAb treatment followed by 200 µg i.p. every 5 days to maintain depletion. Depletion efficiency was confirmed by flow cytometry. Tumor burden was quantified by flow cytometry using the prespecified B cell/tumor gating strategy (including GFP-based identification within the appropriate gate, as indicated), and T cell activation, proliferation, and differentiation states were quantified using predefined gating and/or mean fluorescence intensity (MFI) measurements. Survival curves were compared using the log-rank test. Unless otherwise specified, two-group comparisons were performed using two-sided Wilcoxon rank-sum tests.

### *In vitro* killing assay

CD8+ or CD4+ T cells were isolated from healthy donor PBMCs using Dynabeads Untouched CD8+ or CD4+ T cell Kits, respectively (Invitrogen). CD8+ and CD4+ T cells were activated and differentiated for 5 days under the following conditions: all cells were stimulated with plate-bound anti-CD3 (clone OKT3, Biolegend, 5 μg/mL) and soluble CD28 (clone CD28.2, Biolegend, 1 μg/mL). The following cytokines were added to culture to drive differentiation into distinct T cell subsets, as previously reported: Tc1 (CD8) or Th1 (CD4) (IL-2, 50 IU/mL, IL-12, 5 ng/mL), Tfh (Activin A, 50 ng/mL, IL12, 1 ng/mL, IL-23, 10 ng/mL, TGF-β, 1 ng/mL), Treg (IL-2, 50 IU/mL, TGF-β, 5 ng/mL, sodium butyrate, 100 μM). In some cases, T cell subsets were sorted based on (Tc1, CD8+; Th1, CD4+CXCR6+; Tfh, CD4+PD-1+CXCR5+; Treg, CD4+CD25+TIGIT+) (**Supplementary Figure 5B**). T cells were then co-cultured with RAJI B cells that had been stably transduced with luciferase (Lenti-LucOS, Addgene #22777) at a 10:1 Tcell:B cell ratio. 24 or 48 hours later, cells were lysed, and residual luciferase was measured using a Promega ONE-Glo EX Luciferase Assay System (#E6120). Viability was calculated by normalizing luciferin luminescence to RAJI cells cultured in the absence of T cells.

## Data Availability

All data produced in the present study are available upon reasonable request to the authors.

## ACKNOWLEDGEMENTS

This work was supported by an R35 CA252982 and 2P50CA217694 from the National Cancer Institute (H.G.W.); Congressionally Directed Medical Research Programs (HGW); Sarcoma Center (H.G.W.); two MSK Steven Greenberg Lymphoma Research Awards (S.A.V., L.F.), a Gabrielle’s Angel Foundation Grant (S.A.V.), funding from the Lymphoma Research Foundation (S.A.V., G.S., H.G.W., K.F.), a Japanese Society for the Promotion of Science Overseas Research Fellowship (K.F.), a Blood Cancer United Career Development Grant (L.F.), and funding from Comedy versus Cancer (S.A.V., G.S.), as well as MSKCC’s Core Grant P30 CA008748. We thank Dr. Andrew Steele for his insightful input. This material is based upon work supported by the National Science Foundation Graduate Research Fellowship under Grant No. 2139291 (M.K.).

## Author contributions

Conceptualization: P.B., K.F., G.A.S., L.F., H.G.W., S.A.V. Methodology: P.B., K.F., G.A.S.,L.F., H.G.W., S.A.V. Investigation: P.B., K.F., Y-H.L., Q.G., M.R., M.D.E., A.D., G.A.S., L.F., H.G.W., S.A.V. Analysis: P.B., K.F., J.R., L.M., M.K., S.M., B.G., C.S.L., G.A.S., L.F., H.G.W., S.A.V. Resources: P.B., K.F., G.A.S., L.F., H.G.W., S.A.V. Data Curation: P.B., M.D. W.R.A., G.A.S., L.F., S.A.V. Writing: P.B., S.A.V. Reviewing and editing: P.B., K.F., D.H., G.A.S., L.F., H.G.W., S.A.V. Visualization: P.B., S.A.V. Supervision: B.G., C.L., G.A.S., L.F., H.G.W., S.A.V. Funding acquisition: G.A.S., L.F., H.G.W., S.A.V.

## Data availability

Single cell and spatial transcriptomic data from tumor-bearing mice and patients will be deposited in GEO (accession codes pending). All other data that support the findings of this study are available from the corresponding author upon reasonable request.

## Materials and Correspondence

Please address all correspondences and/or requests for materials to.

## SUPPLEMENTARY FIGURE LEGENDS

**Supplementary Figure 1. Multimodal immune profiling of BsAb therapy in BCL2/Ezh2 lymphoma-bearing mice.**

(A) Dot plot showing canonical marker-gene expression across major cell populations identified by CITE-seq of splenocytes from tumor-bearing mice treated with IgG control or BsAb (n=4/group).

(B) CD4+ and CD8+ T-cell subtype heterogeneity by single-cell scRNA/CITE-seq. UMAP embeddings of intratumoral CD4 (left; n = 3,842 cells) and CD8 (right; n = 9,520 cells) T cells from the disseminated BCL2/Ezh2 lymphoma model, pooled across IgG control- and BsAb-treated mice (n=4 mice/group), colored by T-cell subtype.

(C) Dot plots validating marker-gene expression across CD4+ (left) and CD8+ (right) T-cell subtypes.

(D) Marker-based validation of spectral flow cytometry population calls used in Figure 1D. Heatmap of relative marker expression across 10 FlowSOM meta-clusters, with expression shown relative to the median for each marker, concatenated across treatment conditions and grouped by assigned cell identity.

(E) Splenic tFLp6 tumor burden, measured as the percentage of GFP+ cells among live cells, at baseline and on days 2, 4, and 6 after IgG or BsAb treatment. Each point represents an individual mouse; boxplots show median and IQR. P values were determined using Wilcoxon rank-sum tests; *p<0.05.

(F) Relative abundance of tFL-P6 tumor cells and the indicated immune-cell populations following IgG or BsAb treatment, expressed as log2 fold change relative to day 0. Data are shown as mean ± SEM. For (E–F), n=3 day 0; n=5 day 2 IgG; n=6 day 4 IgG; n=4 day 2 BsAb; n=6 day 4 BsAb; and n=3 day 6 BsAb. No day 6 IgG samples were available because of mortality.

(G) Mean relative composition of six cell populations (tFL-P6, normal B, myeloid, NK, CD8⁺ T, and CD4⁺ T cells) in the spleen, bone marrow, and lymph nodes 4 days after IgG or BsAb treatment (n = 4 mice per arm per tissue). Population proportions were renormalized to 100%.

(H) Abundance of the same 6 cell populations as a percentage of live cells (unnormalized) in the spleen, bone marrow, and lymph nodes 4 days after IgG or BsAb treatment (n = 4 mice/arm per tissue). P values were calculated using two-sided exact Wilcoxon rank-sum tests comparing IgG- and BsAb-treated mice; *p< 0.05.

**Supplementary Figure 2. BsAb treatment induces systemic immune remodeling across tissues.**

(A) Relative marker expression across CD4+ and CD8+ T-cell subsets in the longitudinal spectral-flow dataset. Values represent median expression across samples, normalized to the median expression of each marker across subsets. TCM, central memory; TEM, effector memory; Tfh, T follicular helper; Treg, regulatory T cells.

(B) Mean fluorescence intensity (MFI) of granzyme B, CX3CR1, Ki-67, PD-1, Tim-3, CD39, ICOS, CXCR6, and CD62L in non-naive CD8⁺ T cells across time points (days 0, 2, 4, and 6) following IgG or BsAb treatment. P values were calculated using pairwise Wilcoxon tests with Benjamini–Hochberg correction for multiple comparisons; *p < 0.05, **p < 0.01.

(C) Mean fluorescence intensity (MFI) of the same nine markers in non-naive, non-Treg CD4+ T cells across time points and treatment. P values were calculated using pairwise Wilcoxon tests with Benjamini–Hochberg correction for multiple comparisons; *p< 0.05.

(D) CD8⁺ T-cell subset abundance (naive, TCM, cytotoxic, exhausted, and TEM; % of parent) across days 0, 2, 4, and 6 following IgG or BsAb treatment (n = 3–6/group).

(E) CD4+ T-cell subset abundance (naive, TCM, Tfh, TEM, cytotoxic, and Treg; % of parent) across days 0, 2, 4, and 6 following IgG or BsAb treatment (n = 3–6/group). For (D, E), P values were calculated by exact two-sided Wilcoxon rank-sum permutation tests comparing IgG and BsAb at days 2 and 4, with Benjamini–Hochberg correction; *p< 0.05, **p< 0.01.

(F) UMAP embedding of 60,000 downsampled events, equally pooled across treatment conditions and analyzed using a myeloid-optimized spectral flow cytometry panel (**Supplementary Table 1**), showing major myeloid populations.

(G) Heatmap showing scaled expression of the indicated markers across tFLp6, B, T/NK, and major myeloid populations in pooled samples analyzed using the myeloid-optimized spectral flow cytometry panel (**Supplementary Table 1**).

(H) Time course of seven myeloid populations (Ly6G⁺ PMNs, classical monocytes, macrophages, patrolling monocytes, pDCs, cDC1, and cDC2; % of live cells) across days 0, 2, 4, and 6 following IgG or BsAb treatment (n = 3–6/group; no day 6 IgG group). P values were calculated by two-sided exact Wilcoxon rank-sum tests comparing IgG and BsAb at days 2 and 4, with Benjamini–Hochberg correction; no comparisons reached significance.

I) UMAP embeddings of myeloid cells (n = 5,913; 1,653 IgG, 4,260 BsAb) colored by subtype.

(J) Dot plot of canonical marker gene expression across myeloid subtypes: Ifit1, Ifit3, Isg15, and Irf7 (Mono_IFN); Vcan and Plac8 (Mono_Transitional); Csf1r, Fcgr1, Itgam, Trem2, and Cd9 (Macro_MonoDerived); Mrc1, Cx3cr1, and Fcgr4 (Macro_TRM); C1qa, C1qb, C1qc, and Cd68 (Macro_C1q); Cpa3, Fcer1a, and Ms4a2 (Mast); and S100a8, S100a9, Retnlg, Cxcr2, and Ly6g (Neutrophil and Stressed_Neutrophil).

(K) Hallmark gene set enrichment analysis (GSEA) comparing BsAb versus IgG across all myeloid cell subtypes.

(L) GESA of phagocytosis- and Fc receptor-related pathways in pooled myeloid cells and individual myeloid subsets following BsAb versus IgG treatment. GO gene sets related to phagocytosis, Fc receptor signaling, immunoglobulin/opsonin binding, or antibody-dependent cellular cytotoxicity were selected; pathways with ≥25% of genes represented in the leading edge are shown. Color indicates normalized enrichment score (red, BsAb enriched; blue, IgG enriched), and dot size indicates −log10(FDR).

**Supplementary Figure 3. T cell phenotypes following anti-CD20 and BsAb treatment and CD4/CD8 T-cell interdependence during BsAb therapy.**

(A) Kaplan–Meier survival of tumor-bearing mice treated with IgG control or anti-CD20 antibody on day 7 after tumor transplantation (n=5/group). P value was determined using a log-rank test.

(B) Relative MFI of the indicated markers on splenic T cells (TCRβ+NK1.1−) 4 days after IgG, anti-CD20, or BsAb treatment (IgG, n=5; anti-CD20, n=7; BsAb, n=8). Each point represents an individual mouse; boxplots show median and IQR. Anti-CD20 and BsAb groups were compared using two-sided Wilcoxon rank-sum tests with Benjamini–Hochberg correction across markers; **p<0.01.

(C) T-cell phenotypes following anti-CD20 or BsAb treatment. Relative mean fluorescence intensity (MFI) of ICOS, PD-1, CD39, CX3CR1, Ki-67, granzyme B (GranB), Tim-3, and CD62L on splenic T cells (TCRβ⁺NK1.1⁻) from BCL2/Ezh2 lymphoma-bearing mice 4 days after treatment with IgG control, anti-CD20, or BsAb (IgG, n = 5; anti-CD20, n = 7; BsAb, n = 8 mice). Boxplots show the median and IQR. Anti-CD20 and BsAb groups were compared using two-sided Wilcoxon rank-sum tests with Benjamini–Hochberg correction across all ten measured markers; CD44 and CXCR6 are shown in **Figure 3B**. **p < 0.01. IgG is shown as a reference and was not included in the statistical comparison.

(D) Experimental schema.

(E) Frequencies of CD4+ T cells (top) and CD8+ T cells (bottom) among total T cells 4 days after IgG or BsAb treatment in mice receiving isotype control, CD4-depleting antibody, or CD8-depleting antibody. Each point represents an individual mouse; boxplots show median and IQR (IgG control+IgG, n=10; CD4 depletion+IgG, n=9; CD8 depletion+IgG, n=7; IgG control+BsAb, n=10; CD4 depletion+BsAb, n=10; CD8 depletion+BsAb, n=10).

(F) Kaplan–Meier survival of tFLp6-bearing mice treated with anti-CD4, anti-CD8, or IgG control antibody, followed by IgG treatment (anti-CD4, n = 8; anti-CD8, n = 7; IgG, n = 7). Dashed line indicates treatment administration at day 7 after transplantation. Log-rank test applied; no significant survival differences across groups.

(G) Representative multiplexed immunofluorescence of spleens harvested 4 days after IgG treatment in mice receiving isotype control, CD4 depletion, or CD8 depletion, showing H&E (10×), CD19, hBCL2, CD4, and CD8 staining with composite overlays.

(H) Heatmaps of marker-expression changes in CD4+ T cells following CD8 depletion (left) and CD8+ T cells following CD4 depletion (right), expressed as log2 fold change in MFI following BsAb relative to IgG treatment.

(I) MFI of the indicated markers on splenic non-naive CD8 T cells 4 days after IgG or BsAb treatment in mice receiving isotype control or anti-CD4 antibody (isotype + IgG, n = 10; anti-CD4 + IgG, n = 9; isotype + BsAb, n = 10; anti-CD4 + BsAb, n = 10).

(J) MFI of the indicated markers on splenic non-naive, non-Treg CD4 T cells 4 days after IgG or BsAb treatment in mice receiving isotype control or anti-CD8 antibody (isotype + IgG, n = 10; anti-CD8 + IgG, n = 7; isotype + BsAb, n = 10; anti-CD8 + BsAb, n = 10). For (I, J), points represent individual mice and boxes indicate median and IQR. P values were calculated by two-sided Wilcoxon rank-sum tests with Benjamini–Hochberg correction. *p< 0.05, **p< 0.01, ***p< 0.001, ****p< 0.0001.

**Supplementary Figure 4. Single-cell profiling of intratumoral T-cell states and clonotypes.**

(A) Frequencies of CD4+ T cells among intratumoral T cells before and during BsAb-based treatment, stratified by progressive disease (PD) status. Each point represents one patient sample; boxplots show median and IQR.

(B–C) Dot plots of canonical marker-gene expression across intratumoral CD4+ (B) and CD8+ (C) T-cell states. Dot size indicates the percentage of cells expressing each gene, and color indicates scaled average expression.

(D) Patient-resolved weighted nearest-neighbor UMAP embeddings of intratumoral CD4+ T cells (top) and CD8+ T cells (bottom), colored by transcriptional state. Seven on-treatment biopsies were profiled: four from patients with complete response, two with partial response, and one with progressive disease.

(E) Per-patient Hallmark gene-set enrichment in CD4⁺ and CD8⁺ T cells. Genes were ranked by pseudobulk log-fold change relative to the other six patients and analyzed across MSigDB Hallmark gene sets using fgsea (CR, n = 4; PR, n = 2; PD, n = 1). Shown are gene sets with FDR < 0.01 in the PD biopsy in either lineage. Columns represent individual patients grouped by best response; color indicates normalized enrichment score (NES) and dot size −log10(FDR.

(F) Distribution of expanded clonotypes across CD4+ states (left) and CD8+ states (right) for each patient. Expanded clonotypes were defined as clonotypes represented by more than one cell. CR, complete response; PR, partial response; PD, progressive disease; Tfh, T follicular helper cells; Treg, regulatory T cells.

**Supplementary Figure 5. CXCR6 prioritization and experimental validation.**

(A) Experimental schema for the polarized human T-cell co-culture assays shown in **Figure 5A**. Human T cells were polarized for 5 days into Tc1, Th1, Tfh, or Treg states, sorted, and co-cultured with luciferase-expressing Raji B cells with or without epcoritamab for 48 hours before measurement of tumor-cell viability.

(B) Representative gating strategy used to isolate the polarized Tc1, Th1, Tfh, and Treg populations.

(C) Luciferase-based cytotoxicity assay using the indicated T-cell populations and Raji B cells in the absence of BsAb. Each point represents an independent replicate.

(D) Log2 fold change in MFI of the indicated markers in splenic CD4+ and CD8+ T cells from Cxcr6^−/−^ relative to WT mice 4 days after BsAb treatment (n=6/group).

(E) Frequencies of the indicated splenic CD4+ and CD8+ T-cell subsets in untreated tumor-bearing WT and Cxcr6^−/−^ mice (n=4/group). Each point represents an individual mouse; boxplots show median and IQR. P values were determined using two-sided Wilcoxon rank-sum tests; *p<0.05.

(F) PET-CT scan obtained 12 months after completion of epcoritamab, rituximab, and lenalidomide in the patient shown in **Figure 5H**.

(G) Volcano plot of differential gene expression between CXCR6-expressing and CXCR6-negative intratumoral CD8+ T cells. Dashed lines indicate log2 fold change of 0.5 and adjusted p=0.01. Genes selected for the *CXCR6-associated CD8 effector module* used for **Figure 5H-K** are shown.

**Supplementary Figure 6. Gating strategies for spectral flow cytometry immunophenotyping.**

(A) Representative gating strategy for T-cell immunophenotyping and identification of the indicated T cell populations.

(B) Representative gating strategy for identification of myeloid populations using the myeloid-focused spectral flow cytometry panel.

## SUPPLEMENTARY TABLES

**Supplementary Table 1.** Antibody clones used for spectral flow cytometry.

| Antigen | Clone | Fluorochrome | Manufacturer | Catalog |
| --- | --- | --- | --- | --- |
| **Lymphoid panel** |  |  |  |  |
| CX3CR1 | SA011F11 | APC | BioLegend | 149008 |
| CCR7 | 4B12 | APC/Fire810 | BioLegend | 120119 |
| CD185 (CXCR5) | 2G8 | BB700 | BioLegend | 566492 |
| CD11b | M1/70 | BUV395 | BD Biosciences | 563553 |
| B220 | RA3-6B2 | BUV496 | BD Biosciences | 612950 |
| CD4 | GK1.5 | BUV563 | BD Biosciences | 612923 |
| NK1.1 | PK136 | BUV615 | BD Biosciences | 751111 |
| CD62L | MEL-14 | BV570 | BioLegend | 104433 |
| TCRb | H57-597 | BUV805 | BD Biosciences | 748405 |
| Tim-3 | RMT3-23 | BV421 | BioLegend | 119723 |
| CD8 | 53-6.7 | BV650 | BioLegend | 100741 |
| KLRG1 | 2F1 | BV786 | BD Biosciences | 565477 |
| CD44 | IM7 | BUV737 | BD Biosciences | 612799 |
| CD39 | 24DMS1 | PE | Invitrogen | 12-0391-80 |
| CD279 (PD-1) | RMP1-30 | PE/Cy7 | BioLegend | 109110 |
| CD186 (CXCR6) | SA051D1 | PE/Dazzle594 | BioLegend | 151117 |
| CD20 | GOT214A | BV605 | BD Biosciences | 752751 |
| CD278 (ICOS) | 15F9 | PE/Fire700 | BioLegend | 107715 |
| Ki-67 | B56 | BV480 | BD Biosciences | 566109 |
| Granzyme B | QA16A02 | Alexa 700 | BioLegend | 372222 |
| FoxP3 | FJK-16s | PE/Cy5 | Invitrogen | 15-5773-82 |
| **Myeloid panel** |  |  |  |  |
| B220 | RA3-6B2 | BUV496 | BD Biosciences | 612950 |
| TCR-β | H57-597 | BUV805 | BD Biosciences | 748405 |
| CD11c | N418 | BB515 | BD Biosciences | 565586 |
| CD11b | M1/70 | BUV737 | BD Biosciences | 612800 |
| NK1.1 | PK136 | BUV615 | BD Biosciences | 751111 |
| CD64 (FcγRI) | X54-5/7.1 | BV711 | BioLegend | 139311 |
| Ly-6C | HK1.4 | BV570 | BioLegend | 128030 |
| F4/80 | BM8 | BV605 | BioLegend | 123133 |
| I-A/I-E (MHCII) | M5/114.15.2 | BV650 | BioLegend | 107641 |
| CD163 | S15049I | BV421 | BioLegend | 155309 |
| XCR1 | ZET | BV785 | BioLegend | 148225 |
| PD-L1 (CD274) | 10F.9G2 | PE/Fire640 | BioLegend | 124345 |
| CD39 | Duha59 | PE/Cy7 | BioLegend | 143806 |
| CX3CR1 | SA011F11 | PerCP/Cy5.5 | BioLegend | 149010 |
| Siglec-H | 551 | Alexa Fluor 647 | BioLegend | 129607 |
| Ly-6G | 1A8 | BUV563 | BD Biosciences | 612921 |
| **Lymphoid panel** |  |  |  |  |
| CX3CR1 | SA011F11 | APC | BioLegend | 149008 |
| CCR7 | 4B12 | APC/Fire810 | BioLegend | 120119 |
| CD185 (CXCR5) | 2G8 | BB700 | BioLegend | 566492 |
| CD11b | M1/70 | BUV395 | BD Biosciences | 563553 |
| B220 | RA3-6B2 | BUV496 | BD Biosciences | 612950 |
| CD4 | GK1.5 | BUV563 | BD Biosciences | 612923 |
| NK1.1 | PK136 | BUV615 | BD Biosciences | 751111 |
| CD62L | MEL-14 | BV570 | BioLegend | 104433 |
| TCRb | H57-597 | BUV805 | BD Biosciences | 748405 |
| Tim-3 | RMT3-23 | BV421 | BioLegend | 119723 |
| CD8 | 53-6.7 | BV650 | BioLegend | 100741 |
| KLRG1 | 2F1 | BV786 | BD Biosciences | 565477 |
| CD44 | IM7 | BUV737 | BD Biosciences | 612799 |
| CD39 | 24DMS1 | PE | Invitrogen | 12-0391-80 |
| CD279 (PD-1) | RMP1-30 | PE/Cy7 | BioLegend | 109110 |
| CD186 (CXCR6) | SA051D1 | PE/Dazzle594 | BioLegend | 151117 |
| CD20 | GOT214A | BV605 | BD Biosciences | 752751 |
| CD278 (ICOS) | 15F9 | PE/Fire700 | BioLegend | 107715 |
| Ki-67 | B56 | BV480 | BD Biosciences | 566109 |
| Granzyme B | QA16A02 | Alexa 700 | BioLegend | 372222 |
| FoxP3 | FJK-16s | PE/Cy5 | Invitrogen | 15-5773-82 |
| **Myeloid panel** |  |  |  |  |
| B220 | RA3-6B2 | BUV496 | BD Biosciences | 612950 |
| TCR-β | H57-597 | BUV805 | BD Biosciences | 748405 |
| CD11c | N418 | BB515 | BD Biosciences | 565586 |
| CD11b | M1/70 | BUV737 | BD Biosciences | 612800 |
| NK1.1 | PK136 | BUV615 | BD Biosciences | 751111 |
| CD64 (FcγRI) | X54-5/7.1 | BV711 | BioLegend | 139311 |
| Ly-6C | HK1.4 | BV570 | BioLegend | 128030 |
| F4/80 | BM8 | BV605 | BioLegend | 123133 |
| I-A/I-E (MHCII) | M5/114.15.2 | BV650 | BioLegend | 107641 |
| CD163 | S15049I | BV421 | BioLegend | 155309 |
| XCR1 | ZET | BV785 | BioLegend | 148225 |
| PD-L1 (CD274) | 10F.9G2 | PE/Fire640 | BioLegend | 124345 |
| CD39 | Duha59 | PE/Cy7 | BioLegend | 143806 |
| CX3CR1 | SA011F11 | PerCP/Cy5.5 | BioLegend | 149010 |
| Siglec-H | 551 | Alexa Fluor 647 | BioLegend | 129607 |
| Ly-6G | 1A8 | BUV563 | BD Biosciences | 612921 |

**Supplementary Table 2.**
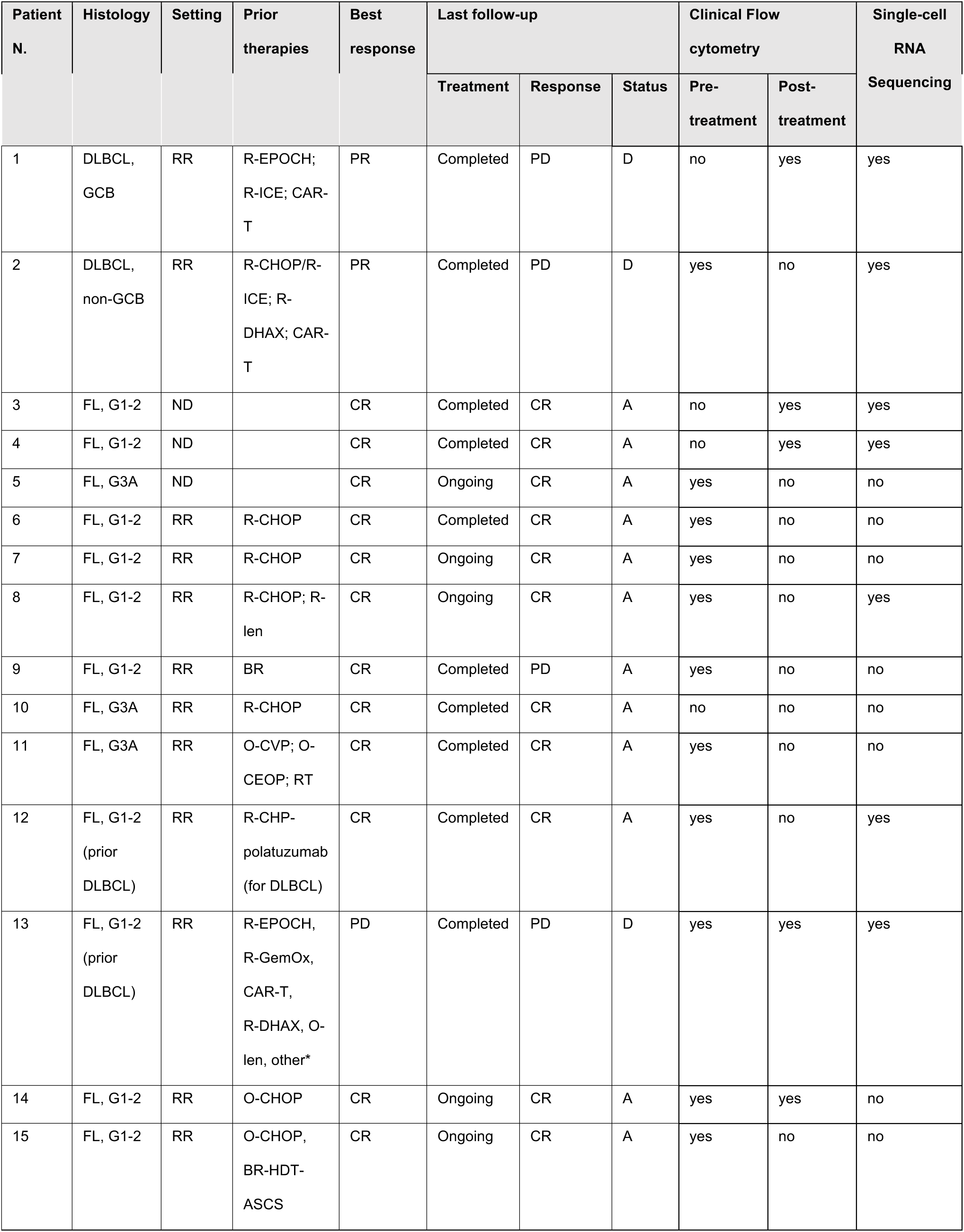
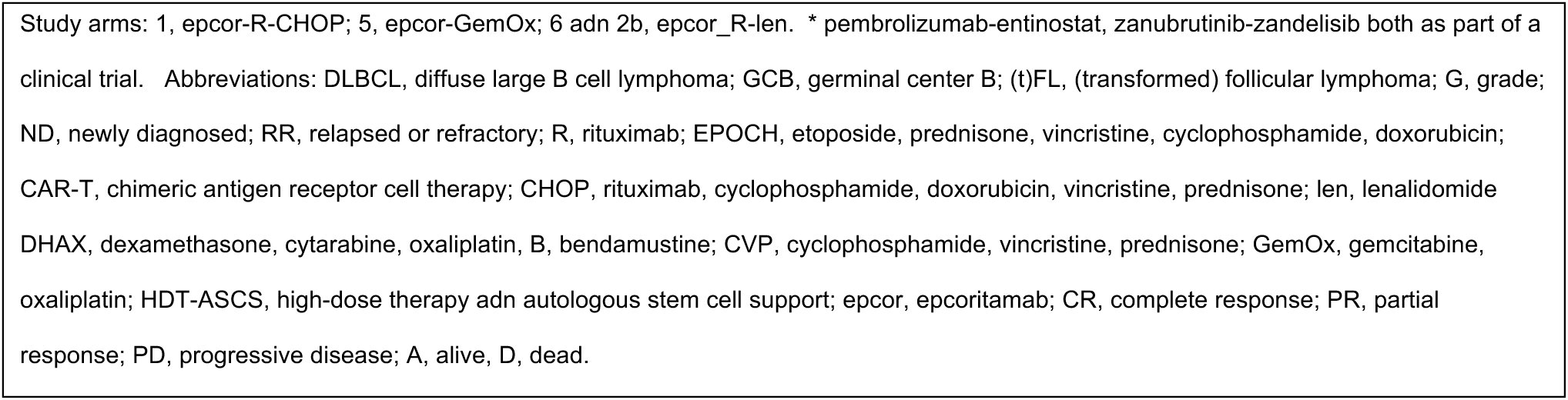
Clinical characteristics and correlative analyses of patients treated on NCT04663347.

**Supplementary Table 3.** Antibody clones used for clinical flow cytometry analysis.

| Antigen | Clone | Fluorochrome | Manufacturer | Catalog |
| --- | --- | --- | --- | --- |
| Kappa | Polytypic | FITC | Agilent | F043401-1 |
| CD8 | REA734 | FITC | Miltenyi | 130-110-815 |
| Lambda | Polytypic | PE | Agilent | R043701-1 |
| CD56 | N901 | PE | Beckman Coulter | IM2073U |
| CD25 | BC96 | PE-Dazzle594 | BioLegend | 302646 |
| CD22 | SJ10.1H11 | PC5.5 | Beckman Coulter | A80712 |
| TCRCβ1 | JOVI.1 | BB700 | BD Biosciences | 7497974 |
| CD19 | J3-119 | PC7 | Beckman Coulter | IM3628U |
| CD10 | ALB1 | APC | Beckman Coulter | IM3633 |
| CD7 | 8H8.1 | APC-Alexa700 | Beckman Coulter | A70201 |
| CD45 | 2D1 | APC-H7 | BD Biosciences | 641408 |
| CD5 | L17F12 | BV421 | Biolegend | 364029 |
| CD38 | HIT2 | BV480 | BD Biosciences | 566137 |
| CD279 | EH12.2H7 | BV605 | BioLegend | 329924 |
| CD20 | 2H7 | BV650 | BD Biosciences | 563780 |
| TCR$\gamma\delta$ | 11F2 | BV711 | BD Biosciences | 624148 |
| CD2 | RPA-2.10 | BV786 | BD Biosciences | 740960 |
| CD26 | M-A261 | BUV395 | BD Biosciences | 744454 |
| CD4 | SK3 | BUV563 | BD Biosciences | 612912 |
| CD3 | UCHT1 | BUV737 | BD Biosciences | 612750 |
| CD14 | M5E2 | BUV805 | BD Biosciences | 612903 |

