## Supplementary Figures for "CXCR6+ T cells are required for CD3xCD20 Bispecific Antibody Efficacy in B cell Lymphomas"

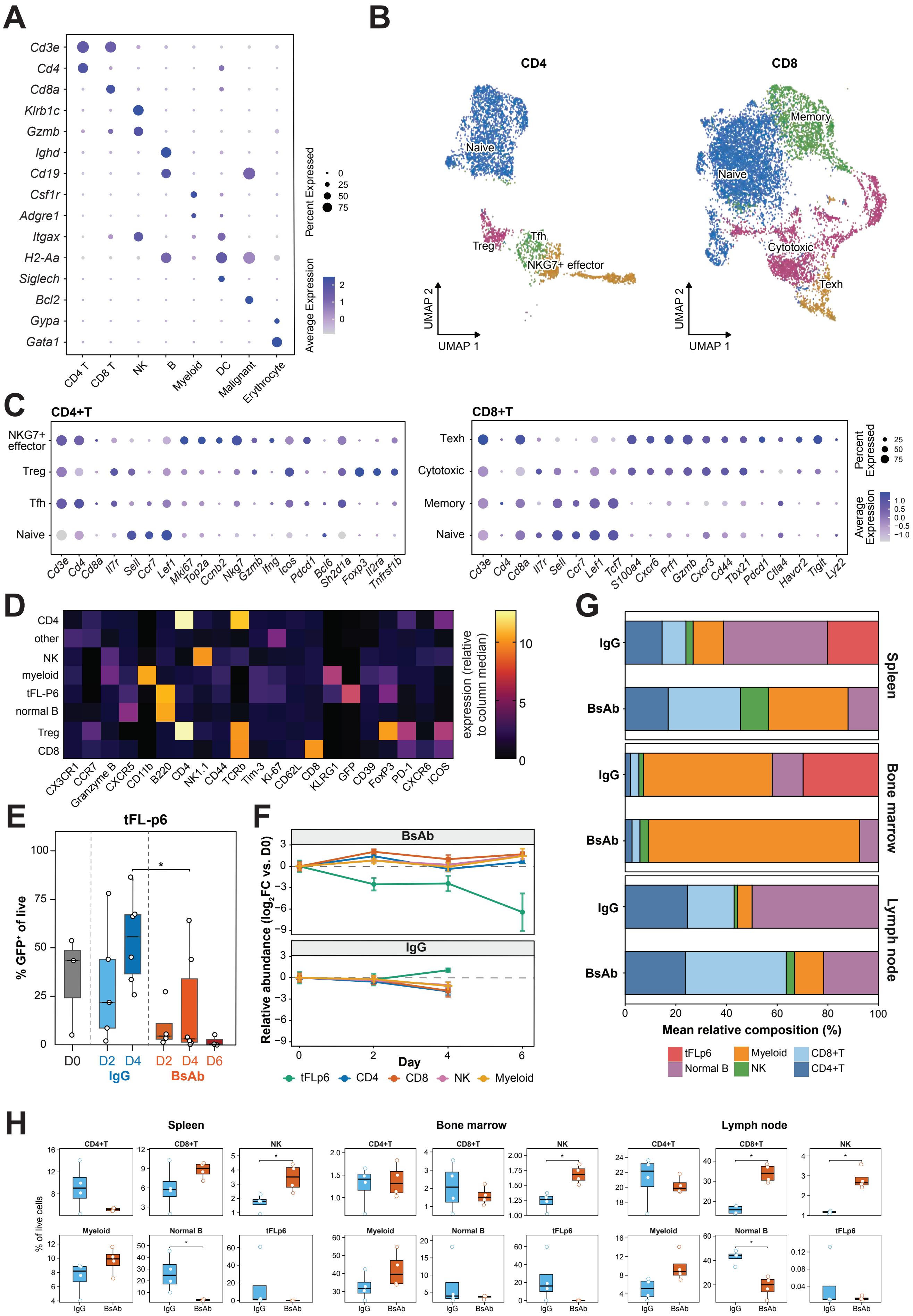


Supplementary Figure 1.

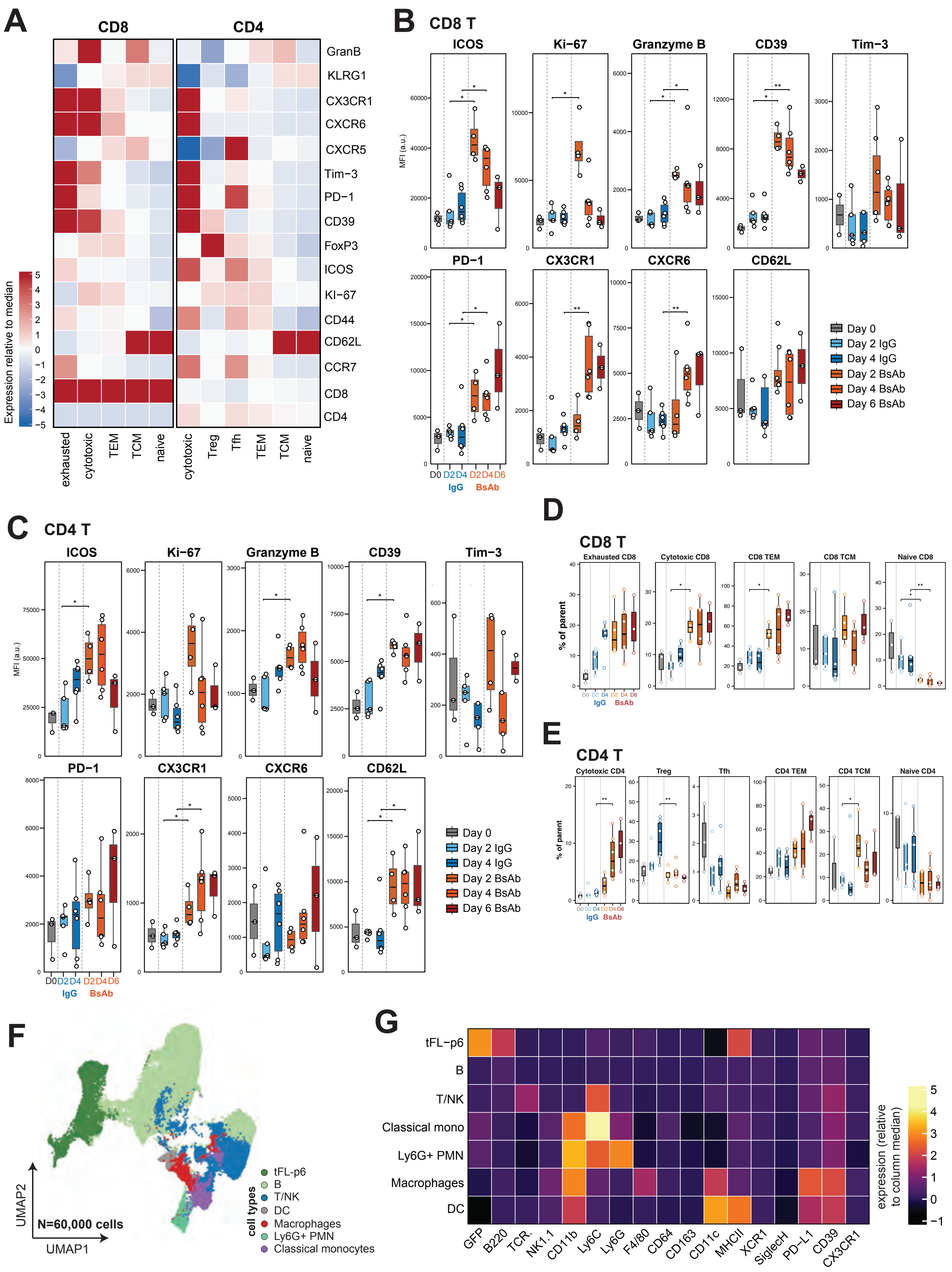


**Supplementary Figure 2.**

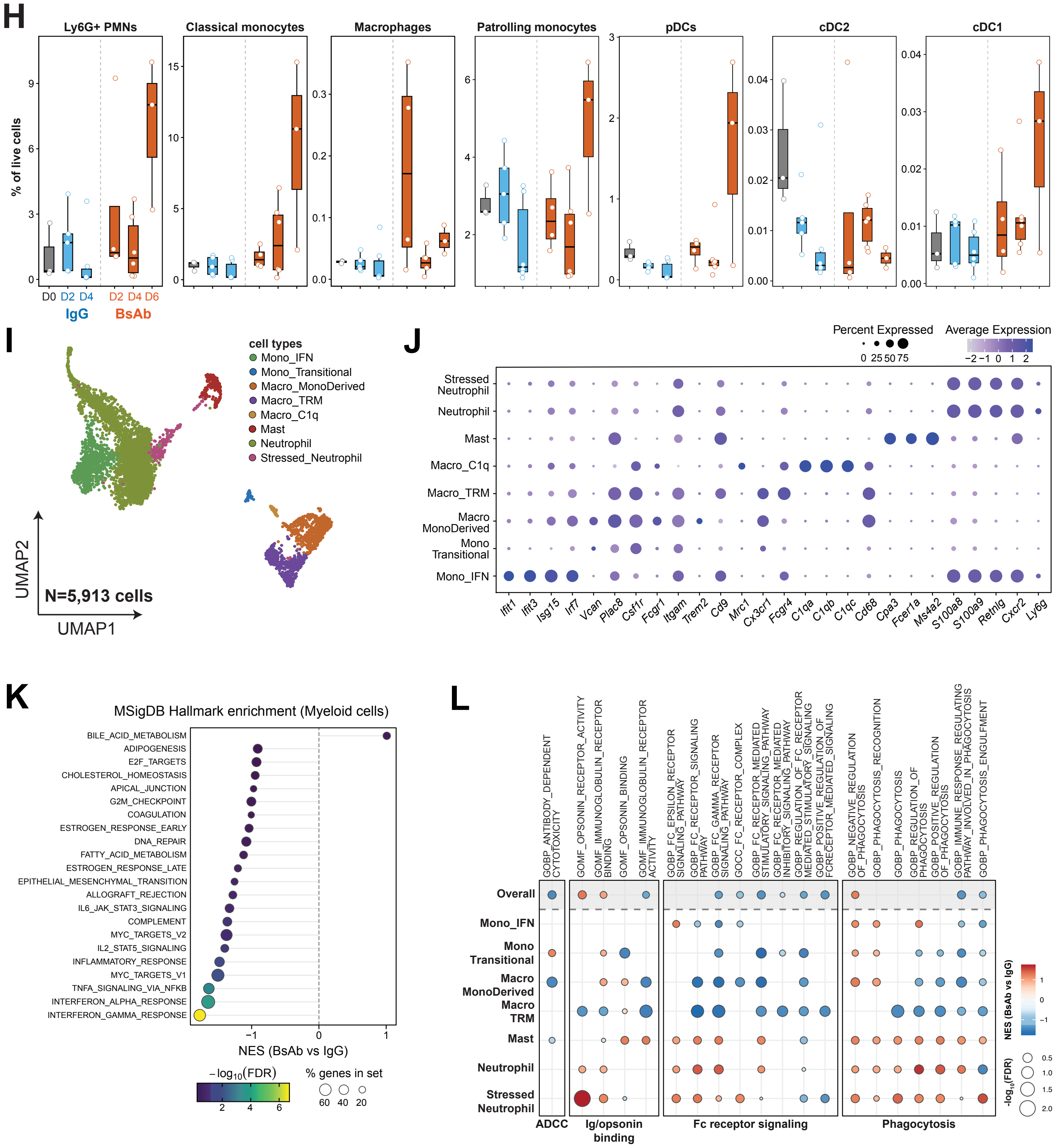


**Supplementary Figure 2** (continued).

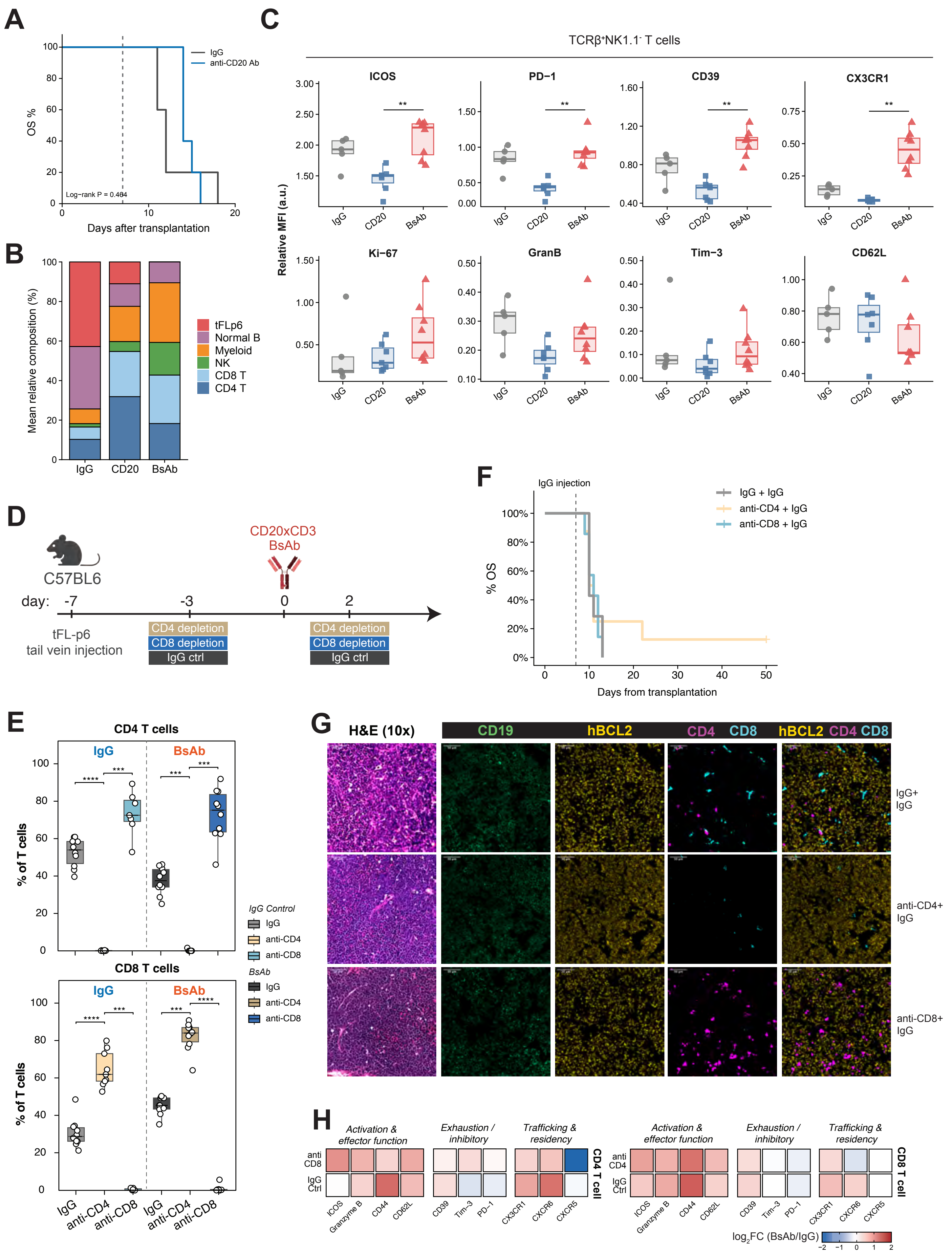


**Supplementary Figure 3.**

I

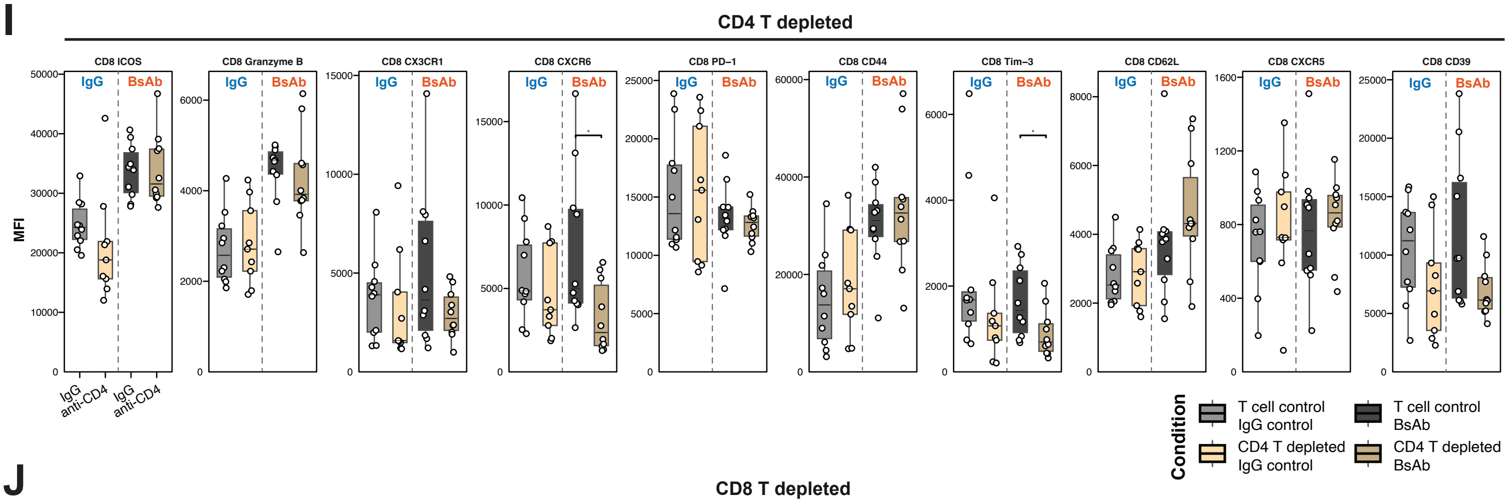


J

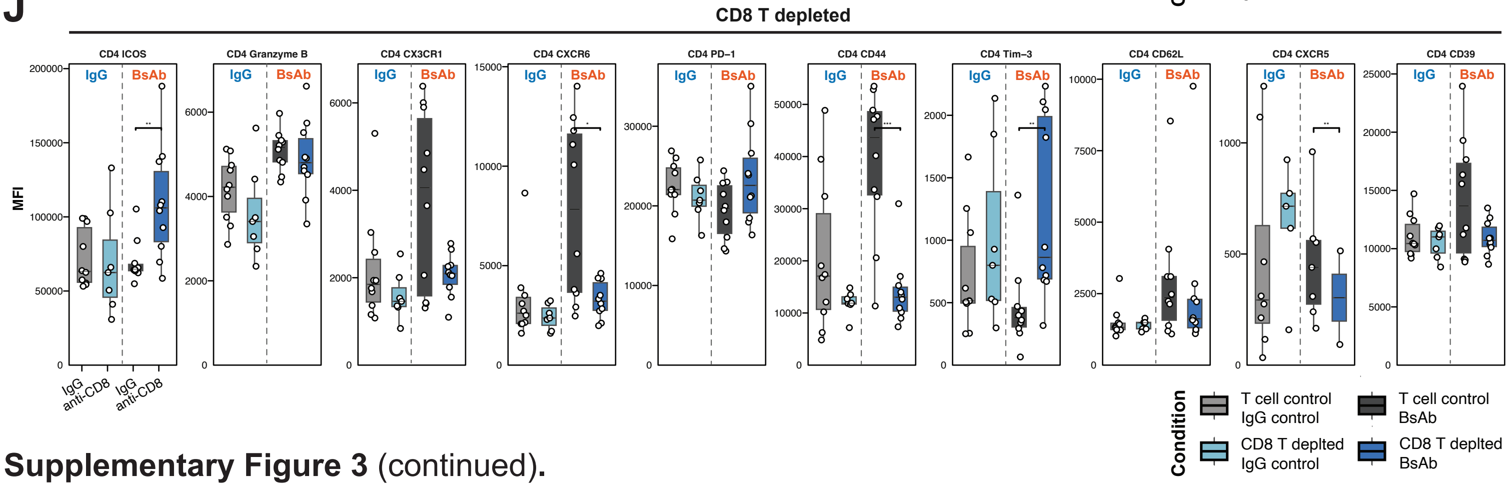


**Supplementary Figure 3** (continued).

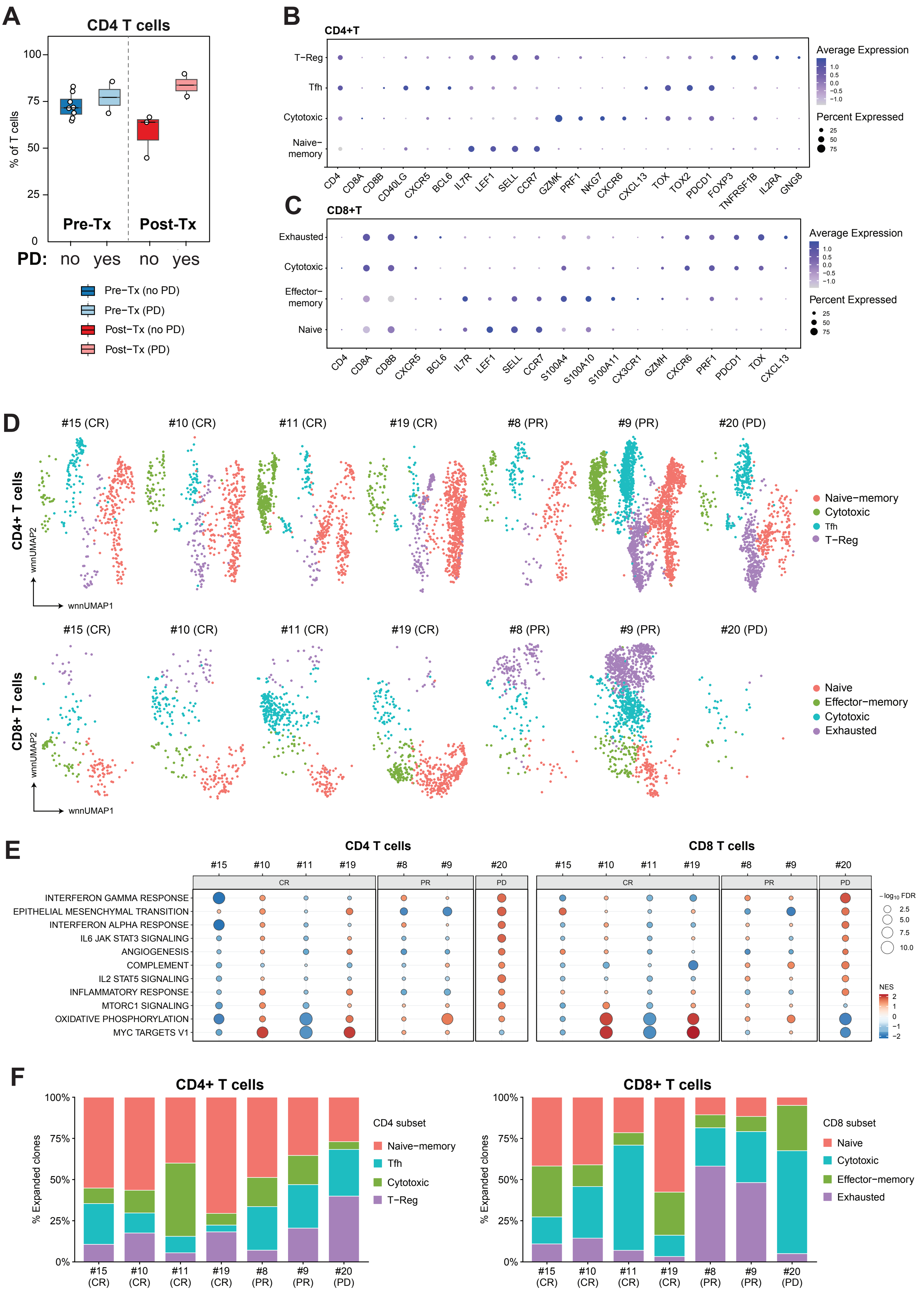


**Supplementary Figure 4.**

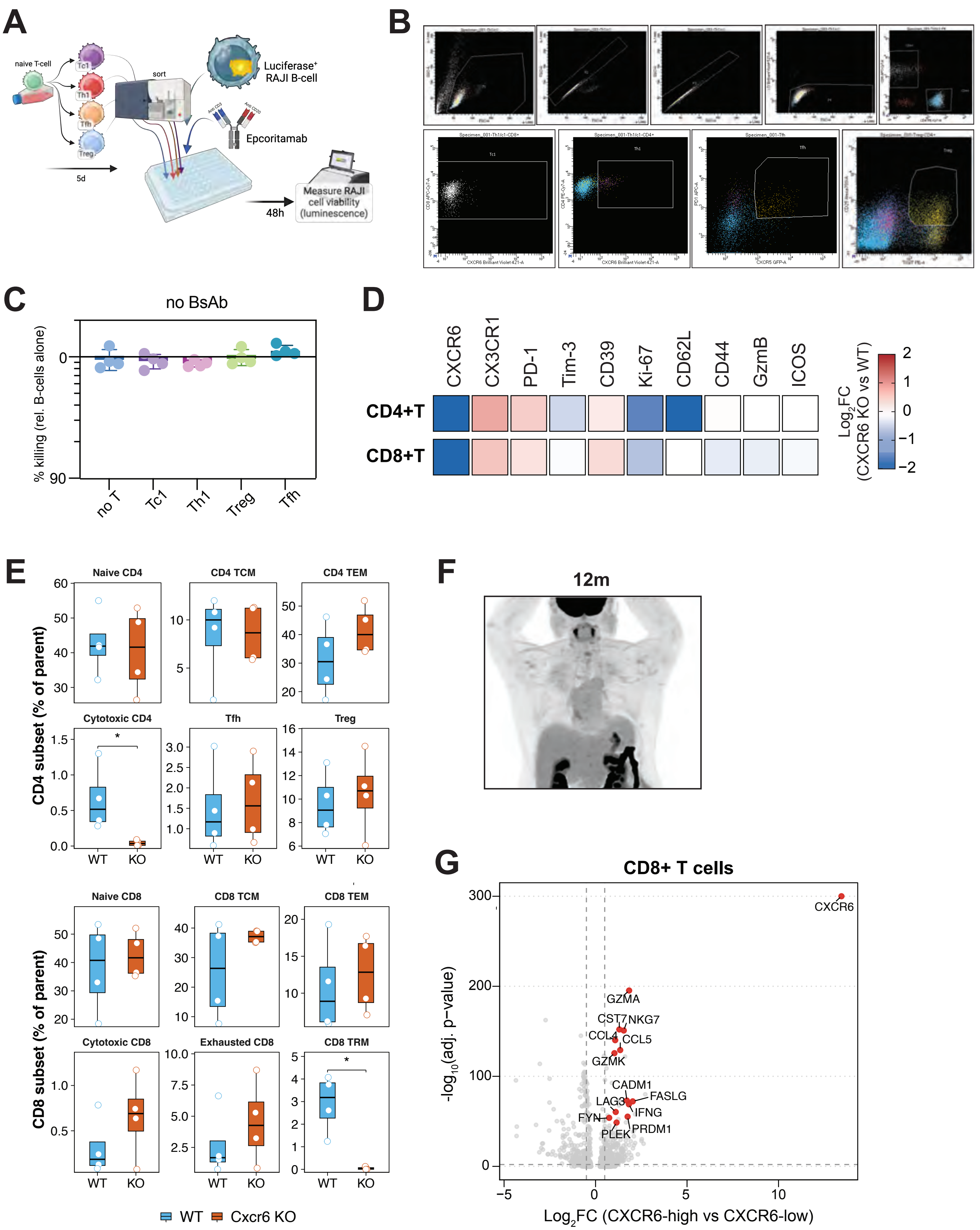


**Supplementary Figure 5.**

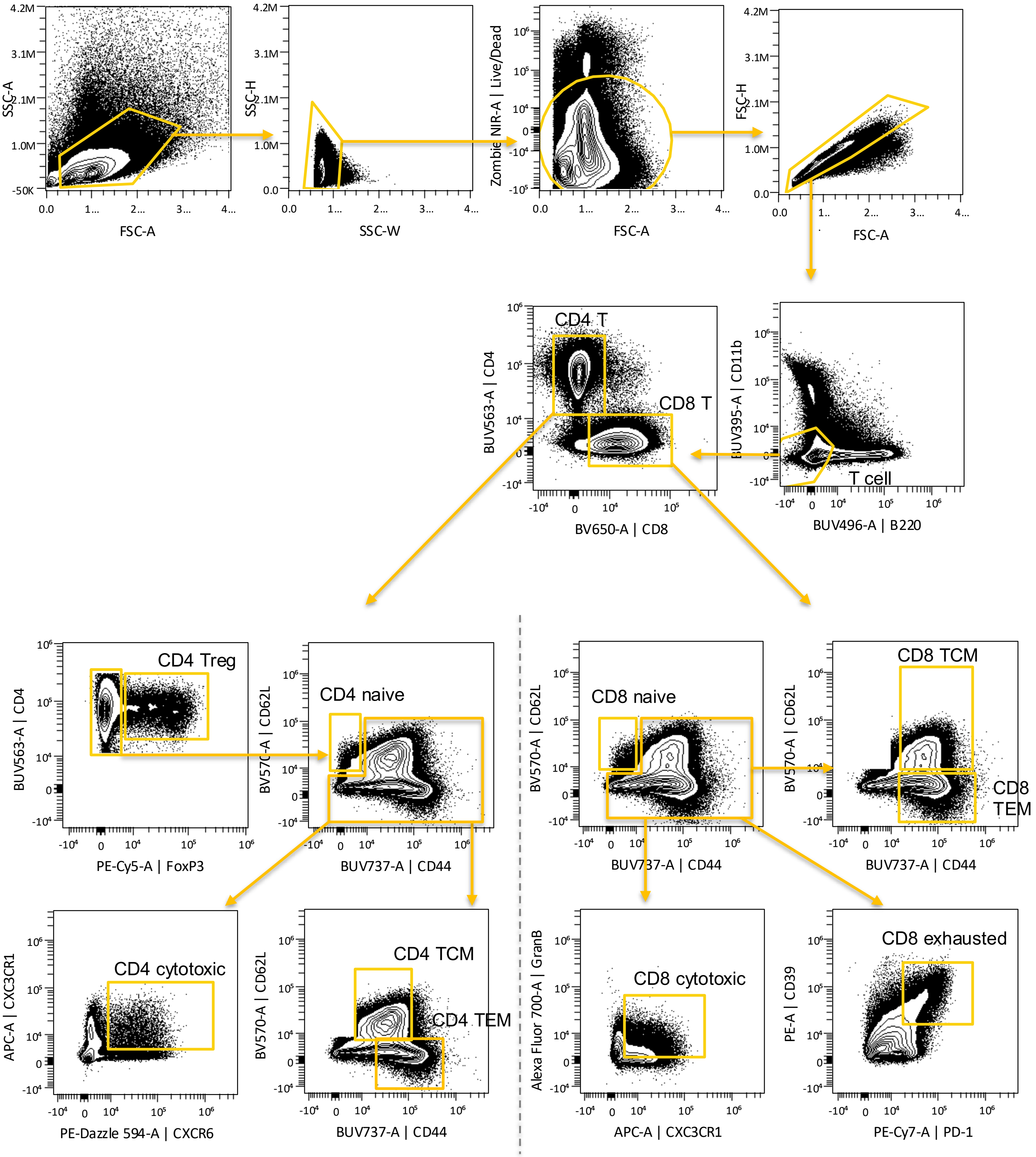


**Supplementary Figure 6.**

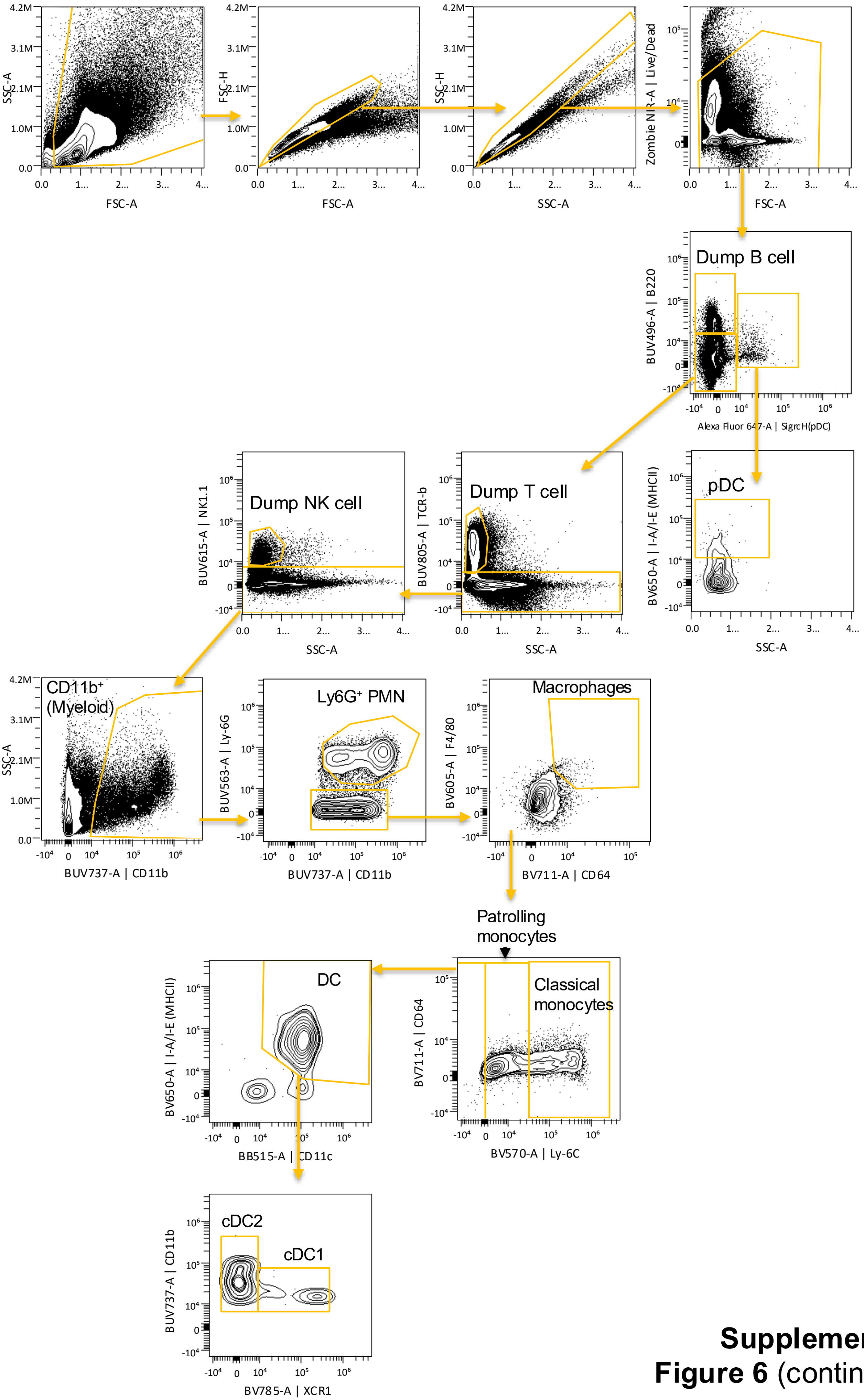


**Supplementary Figure 6** (continued).
